# Cortical Mechanisms of Visual Hypersensitivity in Women at Risk for Chronic Pelvic Pain

**DOI:** 10.1101/2020.12.03.20242032

**Authors:** Matthew J. Kmiecik, Frank F. Tu, Rebecca L. Silton, Katlyn E. Dillane, Genevieve E. Roth, Steven E. Harte, Kevin M. Hellman

**Affiliations:** Department of Ob/Gyn, NorthShore University HealthSystem, Evanston, IL, United States; Department of Ob/Gyn, Pritzker School of Medicine, University of Chicago, Chicago, IL, United States; Department of Psychology, Loyola University Chicago, Chicago, IL, United States; Chronic Pain and Fatigue Research Center, Department of Anesthesiology, University of Michigan, Ann Arbor, MI

**Keywords:** chronic pain, visceral sensitivity, multisensory hypersensitivity, steady-state visual evoked potential (SSVEP), dysmenorrhea

## Abstract

Multisensory hypersensitivity (MSH), which refers to persistent discomfort across sensory modalities, is a risk factor for chronic pain. Developing a better understanding of the neural contributions of disparate sensory systems to MSH may clarify its role in the development of chronic pain. We recruited a cohort of women (*n*=147) enriched with participants with menstrual pain at risk for developing chronic pain. Visual sensitivity was measured using a periodic pattern-reversal stimulus during EEG. Self-reported visual unpleasantness ratings were also recorded. Bladder pain sensitivity was evaluated with an experimental bladder-filling task associated with early clinical symptoms of chronic pelvic pain. Visual stimulation induced unpleasantness was associated with bladder pain and evoked primary visual cortex excitation; however, the relationship between unpleasantness and cortical excitation was moderated by bladder pain. Thus, future studies aimed at reversing the progression of MSH into chronic pain should prioritize targeting of cortical mechanisms responsible for maladaptive sensory input integration.

---

Multisensory hypersensitivity (MSH), which refers to persistent discomfort from multiple sensory pathways, is a common symptom of sensory processing disorders and chronic pain (Bar-Shalita et al., 2019; Bennett, 1999; Ben-Sasson et al., 2009; Molholm et al., 2020). Conceptually, MSH is related to the increased tendency to report somatic symptoms (previously also known as somatization), and is among the foremost risk factors for chronic pain (McBeth et al., 2001). Increased sensitivity to tactile pressure, light, sound, and odors exemplify MSH and are a common phenotype associated with chronic overlapping pain conditions (Geisser et al., 2008; Harte et al., 2016; Hollins et al., 2009; Martenson et al., 2016; Montoya et al., 2006; Schrepf et al., 2018). Some individuals with chronic pelvic pain conditions experience increased sensitivity to visceral input (e.g., uterus, bladder, bowel, rectum) with concomitant mechanical and thermal hypersensitivity in the periphery (Hellman et al., 2020; Larsson et al., 2012; Payne et al., 2017); however, an fMRI study of women with dysmenorrhea reported that alterations in visceral sensitivity of the uterus may not simultaneously impact rectal sensitivity suggesting that generalized MSH is limited (Böttcher et al., 2019). Many have proposed that the “centralized” mechanism of pain sensitization is responsible for MSH (Arendt-Nielsen et al., 2017; Curatolo et al., 2006; Phillips & Clauw, 2011). Nevertheless, it remains unclear whether increased sensitivity is due to sensory pathway hyperexcitability (i.e., efferent/bottom-up) or mechanisms involving higher cortical modulation and integration (i.e., afferent/top-down). Additionally, the extent to which visceral pain is an element of MSH and associated with hypersensitivity in other modalities, such as vision, is unknown. Our understanding of MSH is limited because ideal at-risk cohorts that include evoked responses are rarely studied with sufficient statistical power (i.e., sample size) or temporal precision. A better understanding of the interaction between sensory and integrative processing may inform therapies addressing psychological factors involved in chronic pain conditions, including somatic symptoms (Fishbain et al., 2009) and pain catastrophizing (Galambos et al., 2019; Quartana et al., 2009). For instance, understanding how early-to-late cortical mechanisms, as well as visceral pain, correspond to sensory unpleasantness would suggest brain regions for targeted interventions.

To investigate clinically relevant mechanisms responsible for MSH, we have identified a cohort of women with symptoms of MSH and at-risk for chronic pelvic pain. This cohort of women with dysmenorrhea (episodic menstrual pain) also have increased bladder pain (Hellman et al., 2020) that correlates with clinical symptoms of chronic pelvic pain (Hellman et al., 2018; Tu et al., 2013, 2017). Quantitative sensory testing in this cohort demonstrated widespread reduced pain thresholds and impaired endogenous pain modulation, further indicating increased risk for developing chronic pain (Hellman et al., 2020; Yarnitsky, 2010). To evaluate the visceral contributions to MSH in the present study, we used our validated noninvasive bladder filling task (Tu et al., 2013) and self-reported menstrual pain. We evaluated the visual contributions to MSH by presenting this cohort with an aversive but nonpainful passive visual stimulus while recording scalp electroencephalography (EEG). Participants viewed a rapidly reversing checkerboard pattern presented across increasing intensities of brightness and rated their perceived unpleasantness. Pattern-reversal visual stimuli are known to elicit a steady-state visual evoked potential (SSVEP) evident in the broadband EEG spectra at the presentation frequency (Norcia et al., 2015; Vialatte et al., 2010). SSVEPs are robust measures of cortical excitation to a visual stimulus with sources primarily located in primary visual cortex (V1) and secondary visual cortex sensitive to motion (V5/MT; Di Russo et al., 2007).

Given previous literature linking visual discomfort and cortical measures across various visual stimulus parameters (Haigh et al., 2013; O’Hare, 2016; Patterson Gentile & Aguirre, 2020), we hypothesized that brightness intensity would modulate SSVEP amplitudes, particularly in V1 (electrode Oz). Given the comorbidity of MSH in chronic visceral pain, we hypothesized that the relationship between visual unpleasantness and cortical excitation would be similarly affected by bladder pain. Notably, in other research that did not evaluate bladder pain, menstrual pain and somatic symptoms were among the factors most strongly associated with chronic pelvic pain (Westling et al., 2013). In the present study, we accounted for somatic symptoms and menstrual pain to differentiate the contributions of these common comorbid symptoms to behavioral and neural measures of MSH. Analysis of the relationship of self-reported responses to visceral and visual provocation with cortical recordings allows us to address the following questions: 1) Does presenting increasing brightness intensities of a visual stimulus coincide with evoked cortical excitation? 2) Do women who exhibit visceral pain sensitivity (either menstrual or bladder) report heightened unpleasantness during visual stimulation? 3a) Is visual unpleasantness associated with increased cortical excitation? 3b) If so, is this relationship moderated by visceral sensitivity?

## Method

### Participants

The present investigation was part of a larger clinical trial “Deciphering the Hormonal and Nociceptive Mechanisms Underlying Bladder Pain” (NCT02214550) that enrolled 378 reproductive-age (18-45) women between 2014-2020. Participants were recruited by flyers posted on local college campuses, advertisements on public transportation, and by referral from nearby gynecology clinics. Potential participants who passed an eligibility screening over the phone were scheduled for an initial screening visit. Original inclusion criteria as part of a larger longitudinal trial included controls, dysmenorrhea participants, and chronic pain participants. Control participants had pain ≤ 3/10 with menses on a 0-10 numerical rating scale (NRS, 0 – no pain, 10 – worst pain imaginable) and no concurrent chronic pain diagnoses. Dysmenorrhea participants had self-reported pain > 4/10 with menses on average, ≥ 4 as demonstrated by diary data. Participants with chronic pain conditions other than dysmenorrhea were excluded from these analyses to avoid potential confounding of the many factors (e.g., anxiety, depression, catastrophizing) that are awry in chronic pain (Geisser et al., 1994).

Controls and dysmenorrhea participants were pooled for dimensional analyses following our observation that women with dysmenorrhea often have heightened bladder pain ratings compared to control participants (Tu et al., 2013). This dimensional strategy is recommended to avoid artificial boundaries and artifacts associated with historical diagnostic criteria, increase relevance to preclinical forms of disease, and improve statistical power (Cohen, 1983; Royston et al., 2006; Yee et al., 2015). Exclusion criteria for the present study included: a) presence of active pelvic or abdominal malignancies, b) absence of regular menses, c) active genitourinary infection in the last four weeks, d) unable to read or comprehend the informed consent in English, e) unwilling to undergo pelvic examination/testing, f) presence of hypertension or risk for developing hypertension, g) unwilling to withdraw from oral contraceptives for two months before the study visit, h) inadequate visual acuity to identify 3mm letters on a monitor 1 m away, or i) hairstyles that precluded EEG cap placements.

From the 378 enrolled participants, 93 were classified into groups (e.g., painful bladder syndrome, chronic pain) that are out of scope for the present investigation, 95 did not complete the assessment visit, 40 were excluded due to technical difficulties (e.g., poor EEG quality resulting from equipment malfunction, capping difficulties due to hairstyle, etc.), two declined to participate in the EEG portion due to migraine sensitivity, and one was excluded due to recreational/illicit substance during the testing appointment; therefore, data from a total of 147 women were included in the present investigation. All participants provided written consent and were compensated monetarily for their time. All procedures followed the principles and guidelines stated in the Declaration of Helsinki and were approved by the NorthShore University HealthSystem’s Institutional Review Board.

### Procedure

At an initial screening session, participants completed questionnaire measures encompassing medical, surgical, psychological, and gynecological history. A full list of all questionnaires administered in the larger clinical trial are reported elsewhere (Tu et al., 2020); however, those pertinent in the present investigation are described here and reported in Table 1. In particular, a subset of questions from the Brief Symptom Inventory (BSI; Derogatis & Melisaratos, 1983) representing the somatic symptom subscale evaluated the psychological distress related to the perception of bodily discomfort. Participants rated on a 5-point scale of distress a series of questions that assessed perceptions of bodily pain, such as “Faintness or dizziness”, “Pains in heart or chest”, “Numbness or tingling in parts of your body”, etc. Scores were summed across these questions for each participant and provided a total BSI score, hereafter referred to as “somatic symptoms.”

**Table 1.** Participant Demographics.

| Measure | Response | N | % of N or M (SD) | Range |
| --- | --- | --- | --- | --- |
| Age (years) |  | 147 | 24.1 (6.3) | 18-43 |
| Race | White | 87 | 59.2% |  |
|  | Black or African American | 13 | 8.8% |  |
|  | Asian | 32 | 21.8% |  |
|  | Native Hawaiian or Pacific Islander | 0 | 0.0% |  |
|  | American Indian or Alaskan Native | 1 | 0.7% |  |
|  | Multi-Racial | 13 | 8.8% |  |
|  | Did not respond | 1 | 0.7% |  |
| Ethnicity | Hispanic or Latino | 23 | 15.6% |  |
|  | Not Hispanic or Latino | 124 | 84.4% |  |
| Average number of days bleeding<sup>#</sup> |  | 146 | 5.7 (1.3) | 3-10 |
| <u>Questions for participants with painful periods only:</u> |  | 116 | 78.9% |  |
| Did period pain start at the time of menarche? * | Yes | 60 | 51.7% |  |
| Age (years) that painful periods began (if not at menarche)? |  | 56 | 16.3 (2.9) | 10-28 |
| Years since menarche (first period) without a period? * | Never, always had a regular period | 67 | 57.8% |  |
|  | Less than 1 year | 32 | 27.6% |  |
|  | 1 | 8 | 6.9% |  |
|  | 2 | 3 | 2.6% |  |
|  | 3 | 2 | 1.7% |  |
|  | 5 to 10 | 3 | 2.6% |  |
|  | More than ten years | 1 | 0.9% |  |
| Days of pelvic menstrual pain/month * |  | 116 | 3.7 (2.0) | 1-10 |
| Days of school or work missed in last 3 months * |  | 116 | 2.2 (3.3) | 0-25 |

*Note.* \* participants were administered this question if they answered Yes to painful periods (> 5 out of 10); # one participant was not administered this question because she did not report having a period for the last 6 months due to birth control pill use although she had a period before the EEG visit.

A standardized pelvic exam was performed by a gynecologist (FFT) to identify potential causes of menstrual pain on the first 98 participants (Hellman et al., 2018). Potential clinical exam findings were only observed in eight participants and followed up with ultrasonography. Among these eight participants, three participants had small pelvic cysts (<2.5 x 3.0 cm), and one had subserosal and intramural leiomyomata (<2.5 x 2.5 cm). We discontinued performing pelvic exams to limit potential discomfort and inconvenience, given that most recruited participants had exam profiles consistent with primary dysmenorrhea.

Eligible participants with dysmenorrhea were subsequently scheduled for a mid-luteal phase assessment visit (approximately 17-25 days post-onset of menses). Participants used ovulation tests on days 10-17 to detect luteinizing hormone surges and confirm the menstrual cycle phase (Greenspan et al., 2007). Participants were asked to rate the “average amount of cramping or pain you have experienced during your menstrual period over the past 3 months when not taking any painkillers and on the worst day of your period” using a 0-100 Visual Analog Scale (VAS; 0 – no pain, 100 – worst pain imaginable) (Hjermstad et al., 2011). This question was asked during the luteal phase and thus avoided complications due to variable painkiller use. We confirmed the average intensity of menstrual pain using electronic daily diaries over a full menstrual cycle before the assessment visit (Hellman et al., 2018).

During their mid-luteal phase assessment visit, participants were asked to avoid taking short-acting, over-the-counter analgesics (e.g., ibuprofen, acetaminophen), short-acting opioids (e.g., hydrocodone or oxycodone), and caffeine for at least six hours before arrival. Participants were instructed to also avoid longer-acting over-the-counter analgesics (e.g., naproxen) for at least twelve hours before arrival. We performed comprehensive quantitative sensory testing (Hellman et al., 2020) and noninvasive experimental bladder distension on all participants to assess their bladder pain (Hellman et al., 2018; Tu et al., 2013). Our bladder test mimics clinical retrograde cystometry, starting with an emptied bladder. After oral ingestion of 20 oz of water, participants were instructed to report when they reached three standard levels of bladder urgency: first sensation, first urge to void, and maximal capacity (Abrams et al., 2002). At baseline and each of these time points, we obtained three-dimensional sonographic measurements of the bladder (GE Voluson 750, Wauwatosa, WI), and participants rated their bladder pain and urgency on a 10 cm VAS using a tablet computer. Experimental bladder pain assessment was capped at two hours, even if participants did not reach maximal capacity. Previous investigations from our laboratory have demonstrated that bladder pain ratings at first urge to void is a specific sign of additional bladder pain sensitivity that is observed on retrospective surveys and diaries correlated with clinical markers of bladder pain (Hellman et al., 2018, 2020); therefore, the first urge bladder pain ratings from the bladder distension task, referred to hereafter as “bladder pain”, were used as a moderating factor in regression modeling. After completing this task, we confirmed that bladder pain had returned to baseline levels before beginning the EEG task.

### EEG Instrumentation

Participants were then prepared for EEG recording. Simultaneously, the room was darkened to less than 10 lux ambient light to allow the participant to adapt sufficiently for the visual task. Participants were instrumented with 32 Ag/AgCl active electrodes arranged in the International 10-20 montage (Brain Vision ActiCap). EEG was recorded at 500Hz (1 Hz high-pass and 250 Hz low-pass 20 dB/decade Butterworth filter) using a Brain Vision actiChamp 24-bit A/D amplifier with Pycorder software (BrainVision, NC). Facial and eye movements were recorded from electrodes placed above the right eye, below the left eye, and in the middle of the brow. Electrode impedances were kept below manufacturer guidelines for active electrodes (25 kΩ). The left mastoid served as the online reference, and FPz served as the ground electrode. Participants were instructed to avoid clenching, blinking, speaking, and any facial muscle activity. Standard technical quality inspections were also performed (e.g., requesting participants to blink and verifying signal changes in real-time) throughout the recording. During instrumentation, EEG data were examined to evaluate whether participants were compliant with instructions.

### EEG Experimental Task

We verified sufficient acuity for the visual stimulation task by asking participants to identify eight lines each containing six letters ranging in size from 5 mm to 2 mm. Participants performed the task seated one meter from a 41×30 cm computer monitor with a 100 Hz refresh rate at 23° viewing angle. The same monitor and distance was used for the visual task described below. Stimulus presentation and onset of physiological data collection were controlled by E-prime 2.0 (Psychology Software Tools, PA).

Five blocks of a blue/yellow checkerboard with contrasts (positive and negative flips) alternating at 25 Hz were presented for 20 seconds each (Harte et al., 2016). This 25Hz stimulus frequency was selected because it aligned with the monitor’s refresh rate, produced an irritating flicker, and resided within the frequency range shown to produce robust SSVEPs (Zhu et al., 2010). Each block contained a single maximal brightness intensity (1, 30, 60, 90 or 120 lux) obtained by adjusting the contrast of the image focused on a fixed crosshair centered in the middle of a solid background. Block order was randomized across participants. Participants were instructed to focus on a fixed crosshair centered in the middle of the screen. After each block, participants rated the unpleasantness of the stimuli using the Gracely Box Scale (Gracely & Kwilosz, 1988; Petzke et al., 2005) that lists the numbers 0 to 20 in descending order next to a set of verbal anchors with logarithmically placed validated positions (see Figure 1).

**Figure 1.**
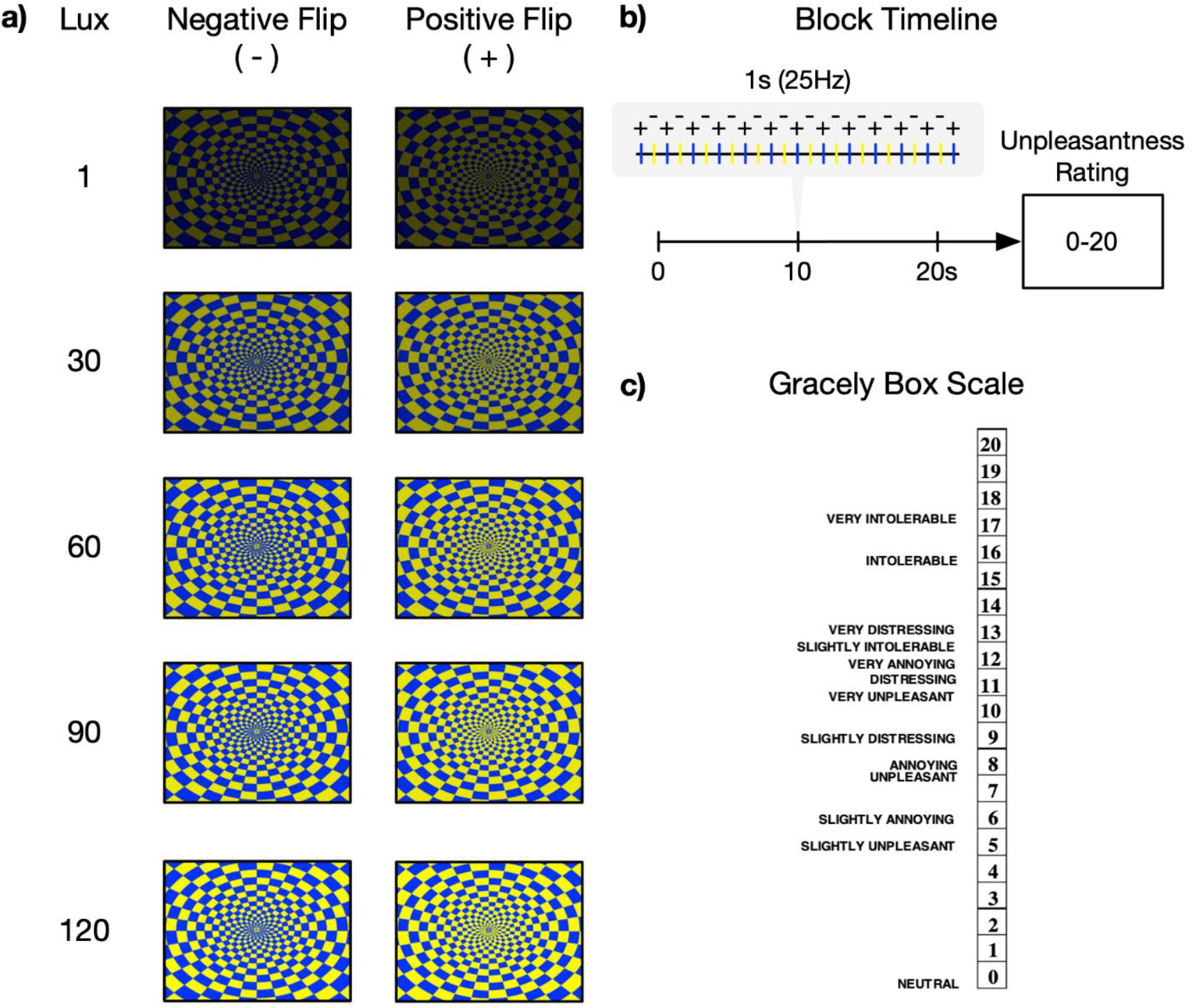
Visual stimulation task presented during EEG recording and designed to elicit an SSVEP. a) Participants viewed an alternating blue-yellow checkerboard pattern with positive and negative reversals across five different intensities of brightness modulated with monotonically increasing lux (i.e., brightness intensity). b) Checkerboards alternated at 25Hz and were presented for 20 seconds before an unpleasantness rating for each brightness intensity. Block order was randomized across participants. c) Participants’ unpleasantness ratings were measured using the Gracely Box Scale with textual descriptors.

### EEG Data Reduction

EEG data were processed in MATLAB using the EEGLAB toolbox (Delorme & Makeig, 2004). EEG data were re-referenced to an average mastoid reference (i.e., the average of the left and right mastoid channels), down-sampled to 256Hz for increased computational efficiency without loss of frequency-based resolution (i.e., Nyquist frequency), and digitally filtered using a 1Hz Hamming windowed sinc finite impulse response (FIR) high-pass filter (−6 dB half-amplitude cutoff, 2Hz transition bandwidth). Line noise (i.e., 60Hz) was removed using the Cleanline EEGLAB plugin (Mullen, 2012). Preprocessed data were then visually inspected and noisy sections of continuous EEG were removed. Clean segments of continuous EEG were submitted to an infomax independent component analysis (ICA) using the “runica” algorithm in EEGLAB (Makeig et al., 1997). Artifactual components were identified (*p* > .6) and removed automatically by the Multiple Artifact Rejection Algorithm (MARA; Winkler et al., 2011) EEGLAB plugin. Continuous EEG was then reconstructed from the remaining independent components and noisy channels spherically interpolated if they maintained uncharacteristic signals throughout the recording (e.g., disconnected channels). Two-second epochs with one-second overlap were extracted from each of the five 20 second stimulation blocks and subjected to artifact rejection using a ±100µV threshold that excluded only 11 epochs (< 1% of eligible epochs). To reduce the spreading of the signal on the scalp due to volume conduction effects and increase topographical specificity, we applied a surface Laplacian spatial filter using the CSD MATLAB Toolbox (Kayser, 2009; Kayser & Tenke, 2006) on the extracted epochs before spectral power calculations that utilized Fast Fourier Transform with Hamming window taper. Power spectral density (PSD) estimates at 25Hz, which was our experimentally controlled visual stimulation frequency, were averaged across the 2-second epochs within each of the five stimulation brightness intensities for the visual task. Therefore, each participant had five PSD estimates used for regression modeling.

## Statistical Analyses

These data were analyzed using multilevel models (MLMs; i.e., linear mixed models) with random intercepts and slopes to model brain-behavior relationships using a model comparison approach (Judd et al., 2017). Clinical studies often differentiate participants into groups (e.g., chronic pain vs. controls) based on metrics with cutoffs that are arbitrary or differ across investigations; however, MLMs provide more flexibility in examining continuous moderating estimates (e.g., bladder pain) that allow for a more sensitive and predictive analysis. The following analyses examined brain-behavior relationships and how these relationships change depending on additional continuous measures, such as pain ratings and somatic symptoms. Importantly, this statistical design accounts for variance at the individual level (i.e., random intercepts and slopes) while estimating between-subject differences, thus providing an optimal method for understanding MSH. All significance testing was two-tailed.

### Analysis 1: Validating the Anticipated Effect of Visual Stimulation on Evoked Brain Activity

Analysis 1 addressed question #1 regarding whether presenting increasing brightness intensities of a visual stimulus coincided with evoked cortical excitation. For each participant and each electrode separately, first level models estimated cortical PSD at 25Hz as a function of linearly increasing brightness:

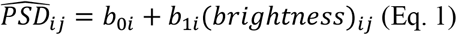

Where *i* refers to an individual participant and *j* refers to brightness intensity. Values for increasing brightness intensity were mean centered: -2, -1, 0, 1, 2. The participants’ regression estimates from first level models were used as dependent variables in second level models that estimated variations in intercepts:

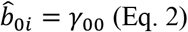

and slopes:

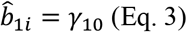

Regression parameters in second levels are designated with γ and subscripts that denote the estimated parameter from level one and level two, respectively. Estimation of the intercept (*γ*_00_) reflects the average PSD across brightness intensities, while estimation of the slope (*γ*_10_) reflects the change in PSD per each increase in brightness intensity.

### Analysis 2: Examining the Relationship Between Visual Unpleasantness and Cortical Excitation with Moderating Factors of Bladder Pain, Somatic Symptoms, and Menstrual Pain

Analysis 2 addressed questions #2, #3a, and #3b listed in the tail of the introduction. Put otherwise, we evaluated the relationships between task evoked cortical excitation and visual unpleasantness and how these relationships were moderated by menstrual pain, bladder pain, and somatic symptoms (see Figure 2). For each participant and each electrode separately, first level models estimated reported unpleasantness as a function of increasing brightness intensity and cortical excitation measured via 25Hz PSD estimates (i.e., SSVEP amplitude):

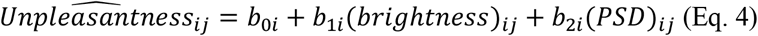

**Figure 2.**
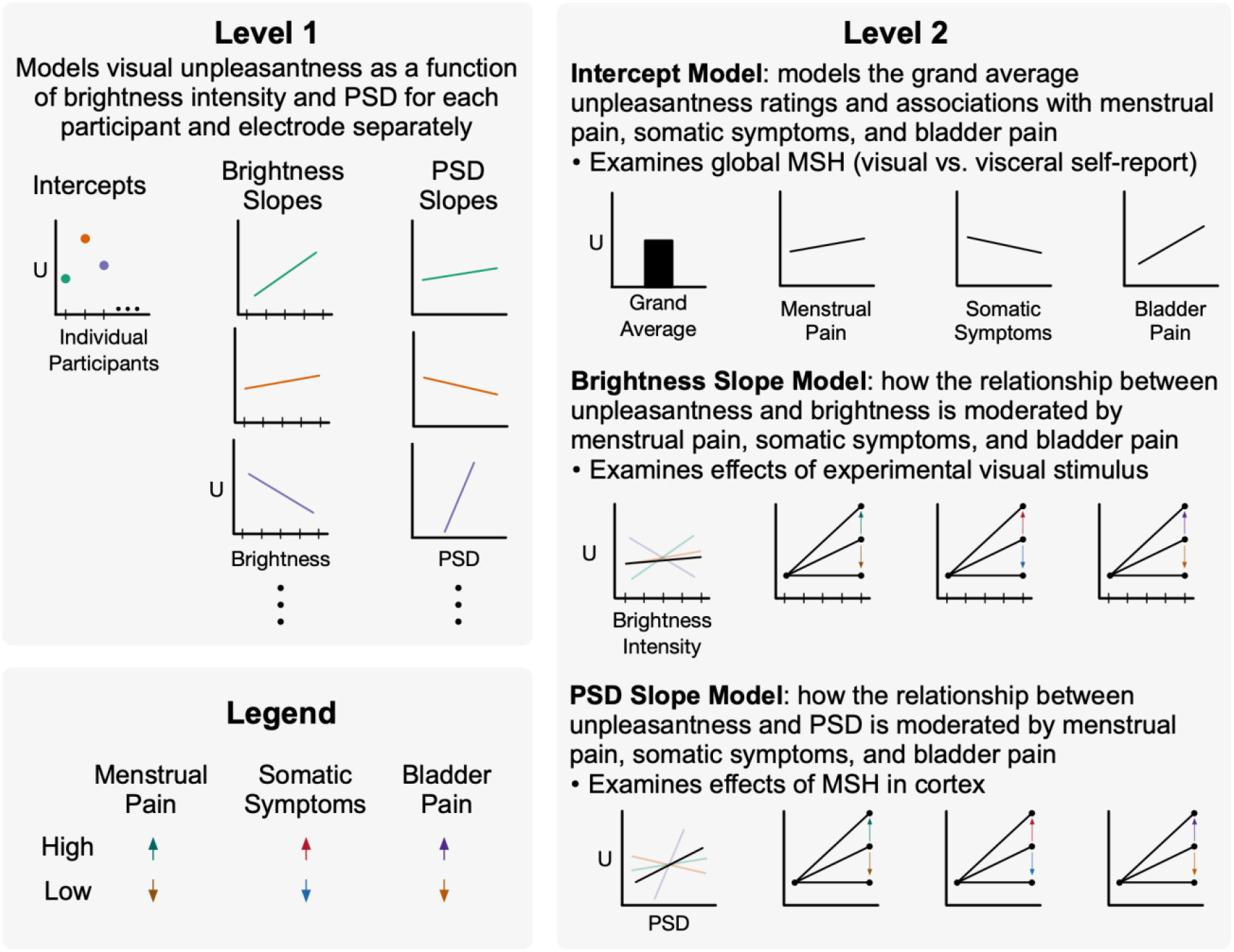
Multilevel modeling allows for comprehensive analysis of MSH accounting menstrual pain, somatic symptoms, bladder pain. In level 1, participant unpleasantness ratings (U) from visual stimulation were modeled as a function of brightness intensity and cortical excitation from 25Hz power spectral density (PSD) estimates. Each participant and electrode were modeled individually, allowing for random intercepts and slopes. Ellipsis indicate all participants are included in this model. Intercepts, brightness slopes, and PSD slopes were modeled separately in level 2 as a function of participants’ prior menstrual pain, somatic symptoms, and bladder pain. This was modeled separately for each electrode but across participants. Moderating effects are demonstrated with arrows depicting high vs. low reported pain/symptoms. Therefore, level 2 brightness and PSD slopes depict positive moderating effects; however, negative moderating effects are also possible (not shown). Data presented are fictional and shown for illustrative purposes. MSH = multisensory hypersensitivity.

Where *i* refers to each participant, while *j* refers to brightness intensity. Both brightness intensity and PSD were mean centered. Regression estimates from first level models were used as dependent variables in second level models that additionally included the mean centered moderating variables of menstrual pain, bladder pain, and somatic symptoms. The first “Intercept Model” estimated moderating effects on the intercept (*b*_0*i*_) that represented average unpleasantness ratings collapsed across brightness intensity:

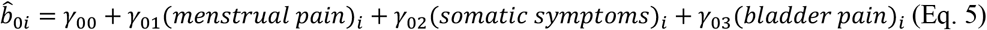

Estimation of the intercept (*γ*_00_) reflected mean unpleasantness ratings at average levels of menstrual pain, somatic symptoms, and bladder pain. Estimation of moderating slopes reflected changes in unpleasantness ratings with changes in menstrual pain (*γ*_01_), somatic symptoms (*γ*_02_), and bladder pain (*γ*_03_). The moderating variables of menstrual and bladder pain addressed question #2. See Supplementary Table 1 for a summary of Analysis 2 regression parameters.

The second “Brightness Slope Model” estimated moderating effects on the brightness slopes (b_1i_) that represented the change in unpleasantness ratings with increases in brightness intensity:

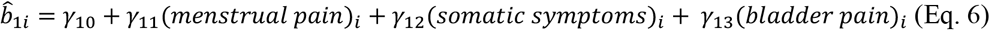

Estimation of the intercept (*γ*_10_) reflected mean brightness slopes at average levels of menstrual pain, somatic symptoms, and bladder pain. Estimation of moderating slopes reflected changes in brightness slopes with changes in menstrual pain (*γ*_11_), somatic symptoms (*γ*_12_), and bladder pain (*γ*_13_).

The third “PSD Slope Model” estimated moderating effects on PSD slopes (b_2i_) that represented the change in unpleasantness ratings with increases in cortical excitation:

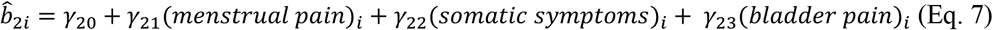

Estimation of the intercept (*γ*_20_), which addressed question #3a, reflected mean PSD slopes at average levels of menstrual pain, somatic symptoms, and bladder pain. Estimation of moderating slopes, which addressed question #3b, reflected changes in PSD slopes with changes in menstrual pain (*γ*_21_), somatic symptoms (*γ*_22_), and bladder pain (*γ*_23_).

We hypothesized that unpleasantness ratings would be modulated by brightness intensity and cortical excitation. Additionally, we hypothesized that these relationships would be moderated by sensory components of MSH, i.e., bladder pain controlling for menstrual pain and non-specific somatic symptoms. We predicted the primary effect of moderation would occur at Oz, given that the neural generators of pattern reversal SSVEPs are located in primary visual cortex (V1) and areas that are sensitive to motion (V5/MT) (Di Russo et al., 2007; Norcia et al., 2015). The remaining 31 electrode sites were exploratory and were corrected for multiple comparisons by controlling for false discovery rate (FDR; Benjamini & Hochberg, 1995). Model assumptions and multicollinearity were examined for violations and deviations from normality.

Statistical analyses were performed in R (version 3.6.3; R Core Team, 2020) and RStudio (version 1.2.5033) using the *dplyr* (Wickham et al., 2020), *broom* (Robinson & Hayes, 2020), and *lmSupport* (Curtin, 2018), and *performance* (Lüdecke et al., 2020) packages, whereas figures were prepared using *ggplot2* (Wickham, 2016), *patchwork* (Pederson, 2019), and *RColorBrewer* (Neuwirth, 2014).

## Results

### Analysis 1 (Question #1): Visual Stimulation Robustly Modulated 25Hz SSVEP PSD

As expected, the visual task effectively elicited a 25Hz SSVEP that increased in PSD with increasing brightness intensities, demonstrating robust stimulation of visual cortex (Figure 3). Specifically, for every increase in brightness intensity we observed an average increase of 2.1 95% CI [1.9, 2.2] dB of 25Hz PSD at Oz (*p* < .001; see Table 2). Put differently, participants’ 25Hz PSD increased an average of 10dB across all brightness intensities (1-120 lux). Although this effect was expected and largest at Oz (partial eta-squared 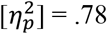, we emphasize that significant linear increases in 25Hz PSD occurred at every electrode site across the scalp even when correcting for multiple comparisons (see Supplementary Table 2). Thus, our visual task effectively evoked widespread cortical excitation precisely synchronized to our stimulus frequency in proportion to stimulus brightness intensity.

**Table 2.**
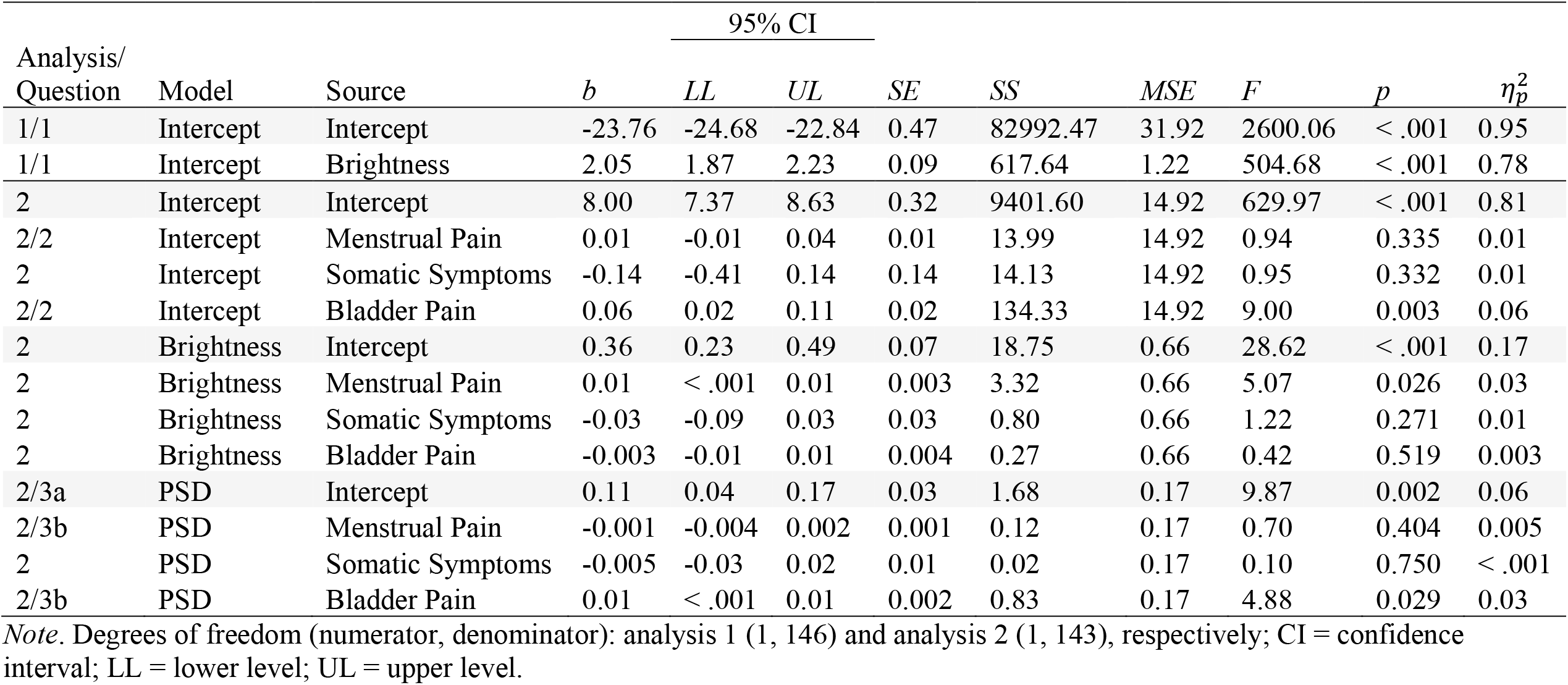
Multilevel Modeling Results at Electrode Oz.

**Figure 3.**
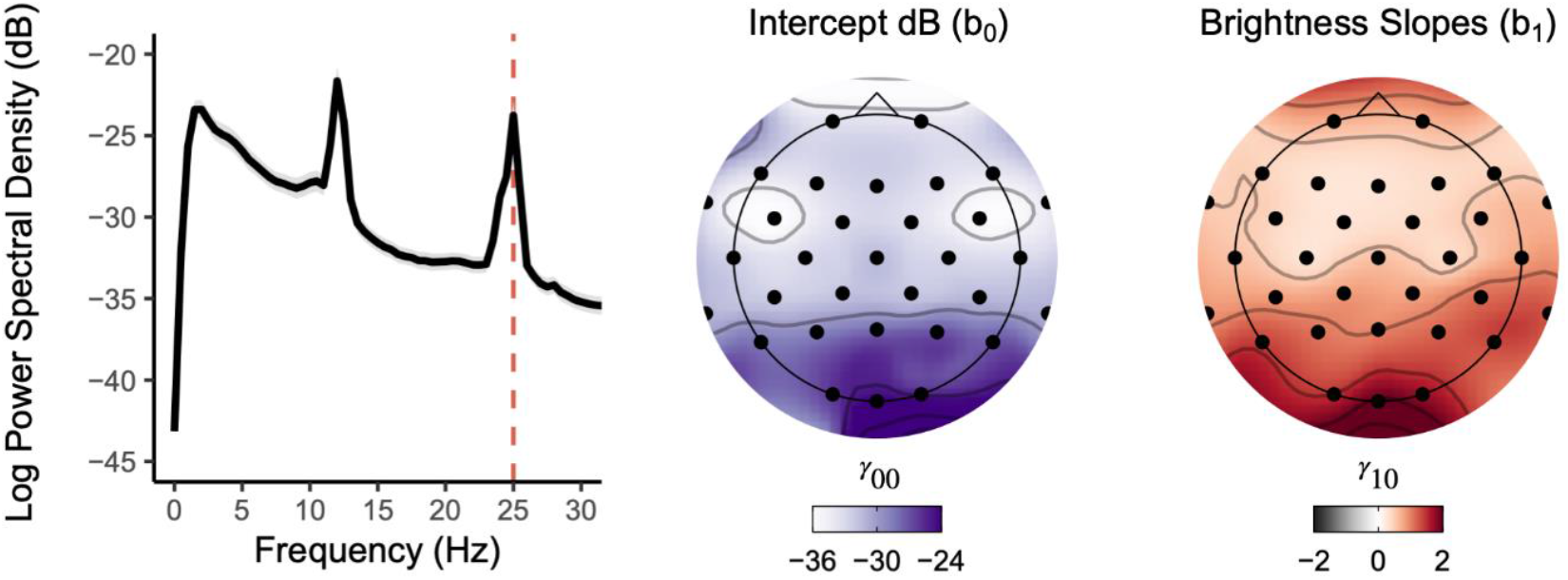
The unpleasant checkerboard stimuli presented at 25Hz evoked widespread robust SSVEPs focused at Oz. *Left*. Broadband PSD averaged across the five presented brightness intensities for all participants shows a clear peak at the 25Hz SSVEP alternating checkerboard frequency. Grey shading denotes 95% confidence interval. *Middle*. Topographically plotted intercepts demonstrated elevated PSD estimates toward occipital electrode sites. *Right*. Topographically plotted regression slopes show scalp-wide positive slopes, especially at occipital sites and Oz, demonstrating an increase in SSVEP PSD estimates with increasing brightness intensities. All topographic sites for intercept and slope effects were significant after correcting for multiple comparisons (*p*_*FDR*_ < .001).

### Analysis 2: Brain-Behavior Relationship Moderated by Prior Menstrual Pain, Somatic Symptoms, and Bladder Pain

The MLM regression procedure modeled brain-behavior relationships and how these relationships were moderated by pain and somatic symptoms. Menstrual pain, somatic symptoms, and bladder pain were positively correlated with each other (see Table 3 for descriptive statistics) and demonstrated variability across their respective ranges (see Supplementary Figure 1).

**Table 3.** Correlations and Descriptive Statistics of Moderating Variables: Somatic Symptoms and Pain.

| Variable 1 | Variable 2 | *r* | Bootstrapped 95% CI |  | *p* | Variable 1 Descriptive Statistics |  |  |  |  |
| --- | --- | --- | --- | --- | --- | --- | --- | --- | --- | --- |
|  |  |  | Lower Level | Upper Level |  | *M* | *SD* | *SEM* | *MIN* | *MAX* |
| Menstrual Pain | Bladder Pain | 0.28 | 0.13 | 0.42 | < .001 | 62.5 | 26.1 | 2.15 | 0 | 100 |
| Bladder Pain | Somatic Symptoms | 0.28 | -0.002 | 0.31 | <.001 | 12.2 | 16 | 1.32 | 0 | 59 |
| Somatic Symptoms | Menstrual Pain | 0.16 | 0.12 | 0.42 | .053 | 2.37 | 2.4 | 0.198 | 0 | 15 |

### Intercept Model (Question #2): How Visual Unpleasantness is Affected by Prior Menstrual Pain, Somatic Symptoms, and Bladder Pain

The intercept model estimated how the participants’ unpleasantness ratings following visual stimulation were moderated by self-reported menstrual pain, somatic symptoms, and bladder pain (Table 2). Collapsing across all brightness intensities, participants rated the unpleasantness associated with visual stimulation as a Gracely Box Scale (GBS) rating of 8.0 [7.4, 8.6] corresponding with the descriptors “annoying” and “unpleasant”. Menstrual pain and somatic symptoms did not moderate mean unpleasantness ratings. However, we observed a positive linear association between bladder pain and mean unpleasantness ratings, suggesting that participants with increased bladder pain rated the visual stimulation as more unpleasant (see Figure 4a). On average, participants’ unpleasantness ratings increased by 1.0 GBS point per 15-point increase in bladder pain 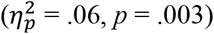.

**Figure 4.**
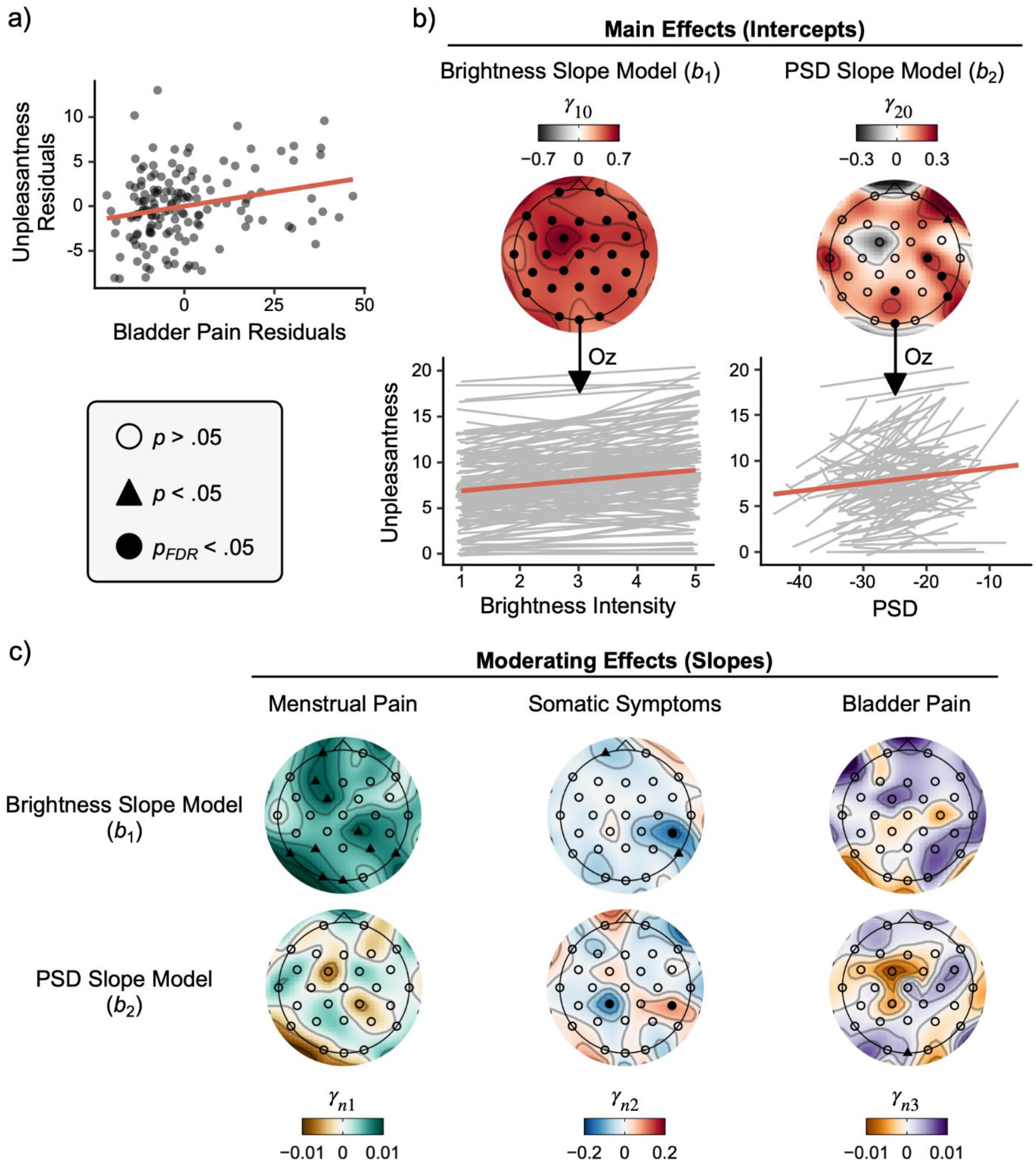
The relationships between visual unpleasantness and brightness intensity/cortical excitation were moderated by menstrual pain, somatic symptoms and bladder pain. a) Partial regression scatter plots depict the positive relationship between bladder pain and participants’ mean unpleasantness ratings averaged across brightness intensities accounting for menstrual pain and somatic symptoms. b) Topographic plots of regression slopes testing the intercepts from the brightness and PSD models. Oz was our *a priori* electrode of interest. Raw (grey) and averaged (red) slopes across all participants plotted below demonstrate that increases in brightness intensity and 25Hz PSD resulted in concomitant increases in participant unpleasantness ratings when accounting for one another. c) Scalp topographies of moderating slopes from second level multilevel modeling results. Given that positive relationships were observed between unpleasantness ratings and brightness/PSD in *b*, positive slopes here depict an increasing positive relationship between moderating variables, while negative slopes depict an increasing negative relationship. Menstrual pain ratings moderated the positive relationship between unpleasantness ratings and brightness, but not 25 Hz PSD at Oz (*a priori* chosen) and several other exploratory electrode sites. Somatic symptoms did not moderate these relationships at Oz; however, somatic symptoms moderated brightness and PSD slopes at a right posterior site (CP6; *p*_*FDR*_ < .05 corrected), despite conflicting directions of moderation. In contrast, bladder pain moderated the positive relationship between unpleasantness and PSD, but not brightness. Level two regression parameter notation *n* denotes both the brightness (*n*=1) and PSD (*n*=2) slope models. PSD = power spectral density; FDR = false discovery rate.

### Brightness Slope Model: How the Association Between Unpleasantness and Brightness Intensity is Affected by Prior Menstrual Pain, Somatic Symptoms, and Bladder Pain

The brightness model estimated whether participants’ slopes predicting unpleasantness as a function of brightness intensity differed significantly from zero on average (i.e., intercepts) and whether this relationship was moderated by self-reported menstrual pain, somatic symptoms, and bladder pain. Given that PSD was estimated alongside brightness in level 1 models (see Eq. 4), brightness slopes are controlled for PSD at each electrode site (i.e., the relationship of unpleasantness and brightness at average PSD). It is for this reason that brightness slopes vary across electrodes because each participant varied in their average PSD for each electrode. After correcting for multiple comparisons, the relationship between unpleasantness ratings and brightness intensity was positive on average across participants at Oz (see Table 2) and across all electrode sites (see Supplementary Table 3), suggesting that participants experienced a monotonic increase in unpleasantness with each increasing brightness intensity (see Figure 4b). More specifically, at average intensities of menstrual pain, bladder pain, and somatic symptoms, the participants reported an increase of .36 [.23, .49] GBS points per increasing intensity level of brightness (*p* < .001), or nearly 2 GBS points from the least to greatest intensity of brightness 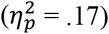.

The relationship between unpleasantness and brightness was moderated by participants’ self-reported menstrual pain at 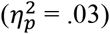 and additional exploratory sites including left posterior, right parietal, and left frontal electrodes 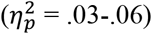. Increases in menstrual pain strengthened the positive relationship (i.e., steeper slopes) between unpleasantness ratings and brightness intensities (see Figure 4c). In other words, participants with the greatest menstrual pain experienced more unpleasantness at equivalent intensities of brightness compared to those with less menstrual pain (see Supplementary Figure 2).

Somatic symptoms moderated the relationship between unpleasantness and brightness at a left frontal electrode (Fp1; 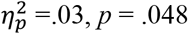) and a right posterior parietal site (P8,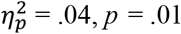). A right posterior parietal site (CP6) was significant even after corrections for multiple comparisons 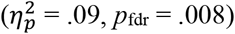. Given its negative slope, this finding suggests that increases in somatic symptoms are associated with an increased negative relationship between unpleasantness ratings and brightness intensities.

Although average unpleasantness ratings were moderated by bladder pain in the intercept model (Figure 5a), the relationship between brightness intensity and unpleasantness was not significantly moderated by bladder pain at any electrode sites (*p* > .2).

**Figure 5.**
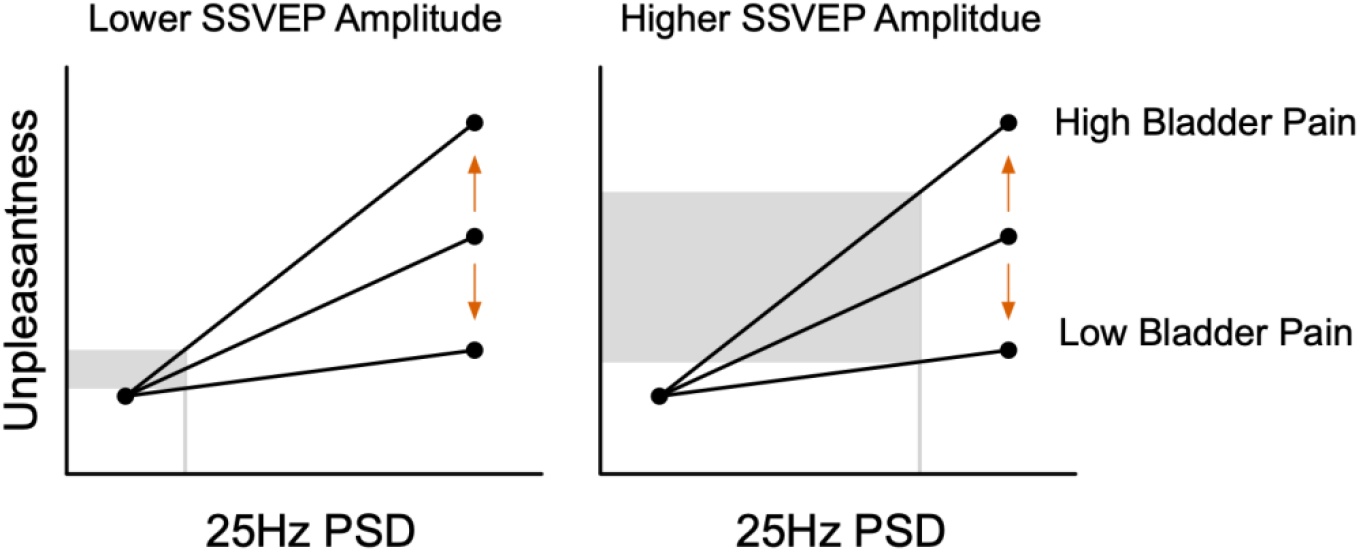
Visceral sensitivity predicts increased visual unpleasantness at equivalent excitation of primary visual cortex. Conceptual line plots demonstrating the moderating effect of bladder pain on the relationship between unpleasantness ratings and cortical excitation measured via 25Hz power spectral density (PSD) estimates (i.e., SSVEP amplitudes) at electrode Oz. This moderating effect implies that when visual cortex is minimally excited by visual stimulation, bladder pain has minimal effect on perceived unpleasantness. However, individuals with greater bladder pain report more unpleasantness when cortical excitation is high. Primary visual cortical excitation is not greater in individuals with heightened bladder pain; rather, downstream interpretation of this signal is likely amplified in women with greater bladder pain.

### PSD Slope Model (Question #3a): The Association Between Cortical Excitation and Unpleasantness

The PSD model estimated unpleasantness as a function of 25Hz PSD (i.e., evoked cortical excitation). Simultaneously, this model also evaluated whether this relationship was moderated by self-reported menstrual pain, somatic symptoms, and experimentally evoked bladder pain. After correcting for multiple comparisons, the relationship between unpleasantness ratings and PSD was positive on average across participants at Oz (see Table 2) and several other central and right parieto-temporal sites (see Supplementary Table 3), suggesting participants experienced a monotonic increase in unpleasantness with increases in 25Hz SSVEP PSD (see Figure 4b). More specifically, the participants reported an increase of .11 [.04, .17] GBS points per increase in 1 dB of 25Hz power at Oz (*p* = .002), or nearly 1.1 GBS points per 10 dB PSD increase in our observed SSVEP 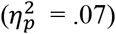. Thus, the relative amount of evoked activity at Oz, even accounting for simulation intensity, is a key predictor of evoked unpleasantness.

### PSD Slope Model (Question #3b): How the Association Between Unpleasantness and Cortical Excitation is Affected by Prior Menstrual Pain, Somatic Symptoms, and Bladder Pain

To determine whether bladder pain specifically moderated the cortical contribution to visual unpleasantness, we examined relationships in a final set of models controlling for menstrual pain and bladder pain (Figure 4c). Although menstrual pain did not moderate this relationship, somatic symptoms did. Somatic symptoms demonstrated conflicting moderating effects on PSD slopes at two parietal electrode sites that survived corrections for multiple comparisons: relationships were both positive (CP6, 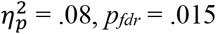) and negative (CP1, 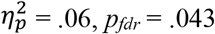). We are cautious about interpreting these effects further given that electrode CP6, an electrode that was not defined *a priori*, negatively moderated brightness slopes (see above) but positively moderated PSD slopes. However, given that electrode CP6 survived multiple comparison corrections in both brightness (see above) and PSD models, we believe this right lateralized effect may represent a cortical phenomenon that differentially responds to brightness and cortical excitation independently of each other.

In the final moderation analysis, we observed that bladder pain positively moderated this relationship, suggesting that women with elevated bladder pain had a stronger positive relationship between unpleasantness and cortical excitation at Oz 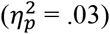. In other words, women with increased bladder pain reported greater unpleasantness ratings at equivalent amounts of cortical response in primary visual cortex (see Supplementary Figure 2).

## Discussion

These findings provide new insights regarding the neural correlates for multisensory hypersensitivity (MSH), which refers to persistent discomfort across multiple sensory modalities, in a cohort of women at heightened risk for chronic pelvic pain (see Supplementary Table 1 for a summary of key findings). Using scalp EEG, we demonstrated that visually presenting increasing brightness intensities of a pattern-reversal checkerboard stimulus evoked increased cortical excitation across the entire scalp, especially at our central occipital site of interest Oz (Question #1). Bladder pain report predicted visual unpleasantness resulting from this stimulus, consistent with the hypothesis that MSH encompasses visceral sensation. We observed a positive relationship between perceived visual unpleasantness and cortical excitation, suggesting that excitation of primary visual cortex, as well as other cortical areas, contributes to increased visual discomfort. Importantly, this relationship was moderated by participants’ rating of bladder pain when accounting for somatic symptoms and menstrual pain; at equivalent primary visual cortex excitation, women with increased visceral sensitivity experienced increased visual discomfort. In other words, the increased visual unpleasantness in individuals exhibiting MSH is not driven by primary visual cortex hyperexcitability. Thus, we theorize that mechanisms in association cortex are responsible for amplifying signals from primary sensory cortex (e.g., visual cortex), resulting in the broad symptoms of MSH and chronic pain vulnerability.

### Bladder Pain Predicts Visual Discomfort (Question #2)

Participants with greater bladder pain rated the visual stimulation as more unpleasant, even after accounting for menstrual pain intensity and somatic symptom profile. This finding demonstrates MSH in this cohort across disparate sensory modalities: visceral and visual. Although visual stimulation is associated with increased unpleasantness in other conditions, like migraine (see Demarquay & Mauguière, 2016) and fibromyalgia (Harte et al., 2016), our study specifically identifies bladder pain as an additional marker correlated with MSH. Menstrual and somatic symptoms did not contribute to visual sensitivity, suggesting that only bladder pain was an indicator of MSH in the present study. Experimental bladder pain is also correlated with the severity of self-reported bladder pain, bowel pain, and intercourse pain in this cohort (Hellman et al., 2020) and in participants with severe chronic pain (Tu et al., 2017). We therefore surmise that bladder pain is a prominent dimension of MSH.

### Cortical Excitation Predicts Visual Discomfort (Question #3a)

We observed a robust brain-behavior relationship that further established the role of cortical excitation in perceived visual unpleasantness. The participants’ perceived unpleasantness to visual stimulation was positively associated with cortical excitation across the entire scalp. Increased unpleasantness was also associated with increased visual stimulation brightness intensity, albeit seen across fewer electrode locations. These findings replicate previous investigations that demonstrated associations between unpleasantness and increased cortical activity in response to aversive visual stimulation (Adjamian et al., 2004; Haigh et al., 2013; Huang et al., 2011; Patterson Gentile & Aguirre, 2020). Our visual task showed that SSVEPs, a measure of cortical excitation, were effectively modulated and related to self-reported measures of unpleasantness, thus promoting the use of SSVEPs conducive to the study of MSH. Although conflicting associations between unpleasantness and cortical excitation were also previously reported (O’Hare, 2017), this discrepancy likely arises due to mechanistic differences in how visual stimulus manipulations, such as brightness and spatial frequency (e.g., gratings), are perceived and interpreted in the brain. Our study supports evidence that increased unpleasantness to a stimulus can be mediated by an increase in sensory pathway excitation (Curatolo et al., 2006), even in a non-somatic pathway such as vision.

### Bladder Pain Moderates the Brain-Behavior Relationship at Oz (Question #3b)

The robust association between visual unpleasantness and cortical excitation was positively moderated by bladder pain. A steeper positive relationship between unpleasantness and cortical excitation was observed in women with greater bladder pain (see Figure 5). This moderation accounted for variance associated with menstrual pain and somatic symptoms, providing evidence that bladder provocation was the predominant factor associated with MSH. Moreover, this effect was observed in our *a priori* electrode of interest Oz, where SSVEP amplitudes are generally observed and likely arise from primary visual cortex (V1) and motion sensitive visual cortex (V5/MT) (Norcia et al., 2015; Vialatte et al., 2010). At average primary visual cortex excitation, women with greater bladder pain experienced more visual discomfort than women with less bladder pain. Otherwise stated, at low cortical excitation there were only slight differences in visual unpleasantness, while these differences were more pronounced at increased brightness intensities, with more unpleasantness experienced by women with greater bladder pain.

This finding implies that primary visual cortical excitation is not greater in individuals with heightened bladder pain; rather, downstream interpretation of this signal is likely amplified in women with worse bladder pain. Harte and colleagues (2016) similarly observed increased insular activation in response to aversive visual stimulation related to pain intensity in fibromyalgia, suggesting the insular cortex as a downstream target for integrative processing and sensory amplification. Lόpez-Solà and colleagues (2014) also observed this amplification in insular cortex and downstream cortical areas; however, concomitant reductions in primary visual and auditory cortex activity were observed in fibromyalgia patients. In chronic pain conditions, altered neural processing in primary cortex, both hypoactivations (López-Solà et al., 2014) and hyperactivations (Lang et al., 2004; Montoya et al., 2006), is commonly observed (Apkarian et al., 2005). We provide contrasting evidence that primary visual cortex is not hyperexcitable in a cohort of women with varying menstrual and bladder pain comorbidity. This discrepancy may emerge from the millisecond temporal resolution of EEG that is methodologically suited to detect the short latency excitation (<50 ms) in primary cortices, unlike fMRI that typically averages signals over several seconds (Ghuman & Martin, 2019).

### Somatic Symptoms Moderate Brain-Behavior Relationships Across Parietal Electrodes

Electrode positions outside our *a priori* region of interest (Oz) did not demonstrate moderation effects between perceived unpleasantness and cortical excitation for either bladder or menstrual pain. Somatic symptoms, however, moderated this association across cortical sites putatively associated with non-specific widespread pain sensitivity. Although many different cortical regions could be responsible for these effects at the observed parietal electrode locations (CP6 and CP1), we can infer potential contributors. The 10-20 system of electrode placement positions CP6 above the angular gyrus (AG) corresponding to Brodmann area 39 (Pascual-Marqui, 1999; Pascual-Marqui et al., 1994). The AG has emerged as a major hub that integrates multisensory information to provide a high-level interpretation of our environment (Seghier, 2013). Structurally, the AG is connected to frontotemporal cortex, medial temporal structures (e.g., hippocampus, parahippocampal gyrus), and the basal ganglia (e.g., caudate nucleus) (Seghier, 2013). Given that the visual task prompts participants to reflect on their experienced unpleasantness during stimulation, it is possible that the AG activity may reflect this downstream contemplative process that integrates multisensory information with past experiences (i.e., medial temporal lobe connections). An example of the AG and its potential multisensory relevance on widespread body symptoms was demonstrated in a study where AG activity was correlated with music-induced analgesia in fibromyalgia patients (Garza-Villarreal et al., 2015). In contrast, electrode CP1 is positioned near the left somatosensory association cortex and is less lateral and more dorsal than the AG. Somatosensory association cortex plays a vital role in deciphering the context of multimodal percepts and emotional processing. For instance, pairing a neutrally valenced visual stimulus with an aversive auditory stimulus resulted in increased negative emotional valence ratings and activation of somatosensory association cortex, even when the visual stimulus was not consciously perceived (Anders et al., 2004). We observed that somatic symptoms moderated relationships in parietal regions in opposite directions across the left and right hemispheres. Neuropsychological models of emotional processing have similarly differentiated valence (pleasant/unpleasant) and arousal (high/low), with unpleasantness and high arousal associated with greater right hemisphere involvement (Heller et al., 1997; Heller & Nitschke, 1998). Given that MSH is a predictor for chronic pain, it would be valuable to establish whether altered activity in parietal cortex also has predictive value.

### Limitations

Some study limitations may qualify our interpretations of the results and generalizability. First, the initial power analysis to plan the broader clinical trial (Hellman et al., 2020) did not account for the present investigation (i.e., *post hoc* aim). Despite this, our sample size (*n* = 147) at an α=.05, was well powered (> 99%) to detect a linear increase in SSVEP amplitude at Oz across brightness intensity 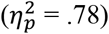 and sufficiently powered (80%) to detect a small-to-medium effect 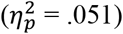. This sample comprised a cohort of women with comorbid menstrual and bladder pain sensitivity that places them at risk for developing chronic pain. These results should be verified in a cohort of severe chronic pain participants and in a comparable risk-enhanced male cohort to assess generalizability across sex.

Second, our study of MSH established a link between specific visceral components (i.e., bladder not uterus) and a disparate non-nociceptive sensory processing modality (i.e., vision). Although we have identified a relationship between bladder pain and other nociceptive modalities, such as pressure and cold pain sensitivity (Hellman et al., 2020), other sensory components were not experimentally measured, such as auditory and olfactory sensitivity. Moreover, our study design and statistical procedure precluded inferences regarding the underlying causes of MSH. Rather, we importantly established brain-behavior relationships between disparate sensory systems that inform mechanisms of MSH and warrant further study. Therefore, future studies would be well served to assess how the presence of visceral sensitivity influences responses at other sensory modalities to broaden our mechanistic understanding of MSH. Third, EEG analyzed with SSVEPs limited our ability to resolve the contribution from subcortical sources (e.g., limbic system, insula), especially given our 32-channel montage. We encourage future studies to employ higher-density EEG methods in conjunction with similar task-evoked steady-state methods to increase source localization specificity.

A key strength of EEG is that it allows for objective temporal measurement of early increased cortical excitation. Our adaptation of this steady-state method, combined with simultaneous behavioral evaluation of unpleasantness, allowed for experimentally controlled electrophysiological measurements of a stimulus intensity-response over a short time (∼2.5 minutes), resulting in a robust small-to-medium effect size 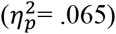. Prior research has mainly relied on questionnaires to evaluate MSH rather than experimental methods (Jones et al., 2006; Yavuz et al., 2013; Zincir et al., 2014), despite the susceptibility of behavioral ratings of discomfort to retrospective biases (Redelmeier & Kahneman, 1996). Given the visual task’s relatively brief administration, other studies can readily extend our findings to clarify pathophysiology through experimental evaluation of brain-behavior relationships in conjunction with questionnaire-based assessments. Given these advantages, the visual task is well suited for simultaneous neuromodulation strategies, such as transcranial magnetic or electric stimulation and neurofeedback (Kayiran et al., 2010; Neeb et al., 2019; Stokes & Lappin, 2010), that seek to reduce MSH and chronic pain.

## Conclusion

Our findings demonstrated MSH in a large cohort of women with comorbid menstrual and bladder pain sensitivity. Brain-behavior relationships between cortical excitation and visual discomfort were moderated by visceral and somatic sensitivity. These results suggest that MSH likely results from maladapted sensory input integration rather than hyperexcitability of sensory afferents in the primary cortex. This evidence emphasizes the need for effective interventions targeting how sensory information is cortically integrated. Mindfulness interventions that decrease general distress also affect top-down modulation (Berkovich-Ohana et al., 2012; Farb et al., 2007; Grant et al., 2011; Jacobs et al., 1996; Travis et al., 2010) and even reduce SSVEP amplitude (Schöne et al., 2018). A future enhanced mindfulness strategy combining simultaneous neurofeedback (Dunham et al., 2019) from the visual task itself could be used to reprogram the neural circuitry responsible for MSH. Similarly, a key next step is determining whether other extant chronic pain treatments—transcranial magnetic stimulation (Grisaru et al., 1998), anticonvulsants (Harte et al., 2016), or antidepressants (Sayar et al., 2005)—could reduce MSH and these associated neural mechanisms in at-risk patients. The paradigm presented here provides a useful experimental method to further evaluate MSH and its neural correlates in order to inform the development of future neuroscience-informed interventions for individuals with chronic pain.

## Data Availability

Data will be made available upon reasonable request and are being currently prepared for a project repository on Open Science Framework (osf.io)

## Data Availability

The data that support the findings of this study are available from the corresponding author upon reasonable request and are currently being prepared for an Open Science Framework repository [link to be posted here upon publication].

## Code Availability

All code used in data processing and analysis of this study are available from the corresponding author upon reasonable request and are currently being prepared for an Open Science Framework repository [link to be posted here upon publication].

## Acknowledgements

The authors thank Dr. G.F. Gebhart for his sagacious advice and editorial assistance, and Ekarin E. Pongpipat for statistical guidance. They are grateful to Ellen Garrison and Nicole Steiner for technical assistance. This study was funded by the National Institute of Child and Human Development (R01HD098193) and the National Institute of Diabetes and Digestive and Kidney Diseases (R01DK100368).

