## Supplementary Material for "Cortical Mechanisms of Visual Hypersensitivity in Women at Risk for Chronic Pelvic Pain"

This document contains the complete supplementary material, including both figures and tables, as referenced in the paper:

Kmiecik, M. J., Tu, F. F., Silton, R. L., Dillane, K. E., Roth, G. E., Harte, S. E., & Hellman, K. M. (2020). Cortical Mechanisms of Visual Hypersensitivity in Women at Risk for Chronic Pelvic Pain. (*full citation appears here*).

Figures and tables begin on the next page, with each display item on its own page.

Supplementary Table 1

*Analysis 2 Regression Coefficient Explanations and Result Summary*

| Parameter | | | | |
|---|---|---|---|---|
| Level One | Level Two | Explanation | Electrode | Result |
| Intercept $b_{0i}$ | Intercept $\gamma_{00}$ | Grand average unpleasantness ratings | All | $M$ = 8, $p$<.001 |
| | Menstrual Pain $\gamma_{01}$ | Association between menstrual pain and unpleasantness | All | *ns* |
| | Somatic Symptoms $\gamma_{02}$ | Association between somatic symptoms and unpleasantness | All | *ns* |
| | Bladder Pain $\gamma_{03}$ | *Association between bladder pain and unpleasantness | All | $\eta_p^2$=.06, $p$=.003 |
| Brightness Slope $b_{1i}$ | Intercept $\gamma_{10}$ | *Association between brightness intensity and unpleasantness | Oz | $\eta_p^2$=.17, $p$<.001 |
| | Menstrual Pain $\gamma_{11}$ | Impact of prior menstrual pain on the association between brightness intensity and unpleasantness | Oz | $\eta_p^2$=.03, $p$=.03 |
| | Somatic Symptoms $\gamma_{12}$ | Impact of prior somatic symptoms on the association between brightness intensity and unpleasantness | Oz | *ns* |
| | | | CP6 | $\eta_p^2$=.09, $p_{fdr}$=.01 |
| | Bladder Pain $\gamma_{13}$ | Impact of prior experimental bladder pain on the association between unpleasantness and brightness intensity | Oz | *ns* |
| PSD Slope $b_{2i}$ | Intercept $\gamma_{20}$ | *How cortical excitability impacts subsequent report of visual unpleasantness | Oz | $\eta_p^2$=.07, $p$=.002 |
| | Menstrual Pain $\gamma_{21}$ | How prior menstrual pain impacts the association between cortical excitability and unpleasantness | Oz | *ns* |
| | Somatic Symptoms $\gamma_{22}$ | How prior somatic symptoms impact the association between cortical excitability and unpleasantness | Oz | *ns* |
| | | | CP1 | $\eta_p^2$=.06, $p_{fdr}$=.04 |
| | | | CP6 | $\eta_p^2$=.08, $p_{fdr}$=.02 |
| | Bladder Pain $\gamma_{23}$ | *How bladder pain affects the association between cortical excitability and unpleasantness | Oz | $\eta_p^2$=.03, $p$=.03 |

*Note*. Unpleasantness refers to ratings of visual unpleasantness during the visual task. ns = not significant; fdr = false discovery rate; * denotes contrasts pertinent to study's primary hypotheses.

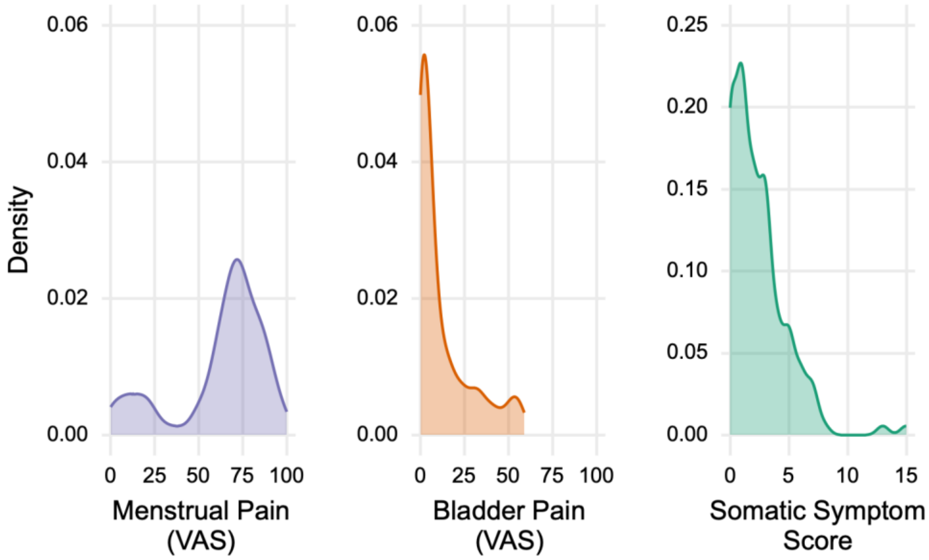


*Supplementary Figure 1*. **Density plots of moderating pain and somatic symptom variables in second level regression models.** The participants had a wide range of menstrual pain, bladder pain, and somatic symptom scores amenable for regression modeling. Y-axis (density) represents the concentration of points about a continuous x-axis measure. VAS = visual analog scale (0 – no pain, 100 – worst pain imaginable).

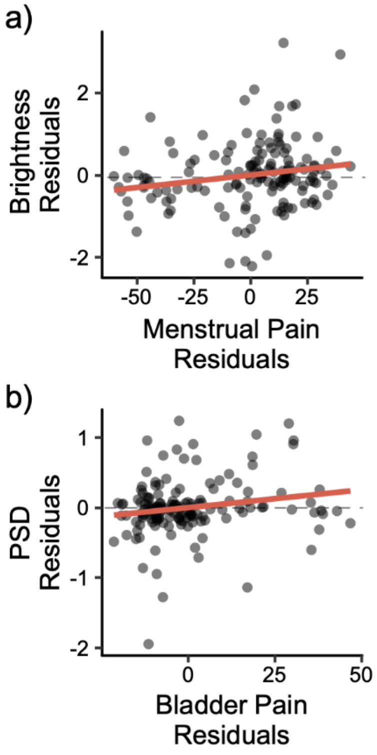


*Supplementary Figure 2.* **Partial regression (added variable) scatter plots depicting the moderating relationship between (a) menstrual pain and brightness slopes [$b_1$] and (b) bladder pain and PSD slopes [$b_2$] at electrode Oz.** These plots depict the unique relationship between the moderating variable (x-axis) and the dependent variable (y-axis) after accounting for the other predictors in the second level models. PSD = power spectral density (25Hz).

Supplementary Table 2

*Analysis 1 Electrode-wise Multilevel Modeling Results: Predicting Power Spectral Density as a Function of Increasing Brightness*

| Electrode | Source | *b* | 95% CI *LL* | 95% CI *UL* | *SE* | *SS* | *MSE* | *F* | $\eta_p^2$ |
|---|---|---|---|---|---|---|---|---|---|
| C3 | Intercept | -33.05 | -33.74 | -32.35 | 0.35 | 160549.52 | 18.14 | 8849.72 | 0.98 |
| C3 | Brightness | 0.30 | 0.21 | 0.40 | 0.05 | 13.56 | 0.35 | 38.94 | 0.21 |
| C4 | Intercept | -32.93 | -33.64 | -32.21 | 0.36 | 159368.39 | 19.08 | 8350.83 | 0.98 |
| C4 | Brightness | 0.40 | 0.29 | 0.51 | 0.05 | 23.71 | 0.43 | 55.05 | 0.27 |
| CP1 | Intercept | -31.87 | -32.39 | -31.35 | 0.26 | 149306.67 | 10.31 | 14478.54 | 0.99 |
| CP1 | Brightness | 0.84 | 0.73 | 0.95 | 0.06 | 103.96 | 0.48 | 217.22 | 0.60 |
| CP2 | Intercept | -32.02 | -32.53 | -31.51 | 0.26 | 150694.86 | 9.81 | 15358.13 | 0.99 |
| CP2 | Brightness | 0.79 | 0.68 | 0.89 | 0.05 | 90.88 | 0.42 | 215.55 | 0.60 |
| CP5 | Intercept | -32.39 | -33.03 | -31.74 | 0.32 | 154180.99 | 15.52 | 9934.66 | 0.99 |
| CP5 | Brightness | 0.82 | 0.68 | 0.95 | 0.07 | 98.07 | 0.67 | 145.77 | 0.50 |
| CP6 | Intercept | -31.35 | -32.10 | -30.59 | 0.38 | 144443.06 | 21.31 | 6777.57 | 0.98 |
| CP6 | Brightness | 1.13 | 0.99 | 1.27 | 0.07 | 187.01 | 0.74 | 252.54 | 0.63 |
| Cz | Intercept | -31.58 | -32.14 | -31.02 | 0.28 | 146617.93 | 11.77 | 12453.33 | 0.99 |
| Cz | Brightness | 0.59 | 0.47 | 0.70 | 0.06 | 50.42 | 0.47 | 107.91 | 0.42 |
| F3 | Intercept | -33.44 | -34.10 | -32.78 | 0.33 | 164362.99 | 16.48 | 9971.85 | 0.99 |
| F3 | Brightness | 0.28 | 0.19 | 0.37 | 0.04 | 11.52 | 0.27 | 42.04 | 0.22 |
| F4 | Intercept | -33.55 | -34.20 | -32.90 | 0.33 | 165465.80 | 16.06 | 10305.60 | 0.99 |
| F4 | Brightness | 0.37 | 0.28 | 0.47 | 0.05 | 20.66 | 0.31 | 67.19 | 0.32 |
| F7 | Intercept | -33.60 | -34.21 | -32.99 | 0.31 | 165957.25 | 13.81 | 12021.46 | 0.99 |
| F7 | Brightness | 0.46 | 0.36 | 0.56 | 0.05 | 30.84 | 0.37 | 82.58 | 0.36 |
| F8 | Intercept | -33.94 | -34.52 | -33.36 | 0.29 | 169314.79 | 12.64 | 13394.94 | 0.99 |
| F8 | Brightness | 0.45 | 0.36 | 0.54 | 0.05 | 30.15 | 0.30 | 98.95 | 0.40 |

| | | | | | | | | | |
|---|---|---|---|---|---|---|---|---|---|
| FC1 | Intercept | -33.15 | -33.64 | -32.67 | 0.25 | 161586.36 | 8.96 | 18026.62 | 0.99 |
| FC1 | Brightness | 0.30 | 0.21 | 0.38 | 0.04 | 13.00 | 0.27 | 48.78 | 0.25 |
| FC2 | Intercept | -33.42 | -33.98 | -32.87 | 0.28 | 164216.73 | 11.70 | 14037.12 | 0.99 |
| FC2 | Brightness | 0.38 | 0.30 | 0.47 | 0.04 | 21.32 | 0.27 | 77.72 | 0.35 |
| FC5 | Intercept | -36.39 | -37.06 | -35.72 | 0.34 | 194656.75 | 16.69 | 11666.25 | 0.99 |
| FC5 | Brightness | 0.43 | 0.33 | 0.52 | 0.05 | 26.58 | 0.31 | 86.56 | 0.37 |
| FC6 | Intercept | -35.96 | -36.56 | -35.36 | 0.30 | 190124.92 | 13.55 | 14028.22 | 0.99 |
| FC6 | Brightness | 0.57 | 0.47 | 0.67 | 0.05 | 47.46 | 0.39 | 120.74 | 0.45 |
| Fp1 | Intercept | -33.59 | -34.28 | -32.91 | 0.34 | 165899.40 | 17.46 | 9500.04 | 0.98 |
| Fp1 | Brightness | 0.71 | 0.59 | 0.83 | 0.06 | 73.90 | 0.56 | 132.13 | 0.48 |
| Fp2 | Intercept | -33.85 | -34.51 | -33.19 | 0.33 | 168419.31 | 16.37 | 10290.52 | 0.99 |
| Fp2 | Brightness | 0.69 | 0.56 | 0.83 | 0.07 | 70.26 | 0.68 | 103.83 | 0.42 |
| FT10 | Intercept | -34.05 | -34.57 | -33.53 | 0.26 | 170412.26 | 10.19 | 16727.84 | 0.99 |
| FT10 | Brightness | 0.61 | 0.49 | 0.73 | 0.06 | 54.89 | 0.51 | 108.27 | 0.43 |
| FT9 | Intercept | -33.19 | -33.69 | -32.69 | 0.26 | 161930.75 | 9.58 | 16900.83 | 0.99 |
| FT9 | Brightness | 0.50 | 0.40 | 0.60 | 0.05 | 36.83 | 0.39 | 94.05 | 0.39 |
| Fz | Intercept | -32.53 | -32.99 | -32.06 | 0.23 | 155525.37 | 8.06 | 19300.29 | 0.99 |
| Fz | Brightness | 0.29 | 0.21 | 0.37 | 0.04 | 12.23 | 0.24 | 50.19 | 0.26 |
| O1 | Intercept | -26.04 | -26.83 | -25.25 | 0.40 | 99674.02 | 23.37 | 4264.77 | 0.97 |
| O1 | Brightness | 1.51 | 1.35 | 1.66 | 0.08 | 333.19 | 0.86 | 386.08 | 0.73 |
| O2 | Intercept | -25.24 | -26.19 | -24.28 | 0.48 | 93641.62 | 34.36 | 2725.20 | 0.95 |
| O2 | Brightness | 1.74 | 1.57 | 1.91 | 0.09 | 446.71 | 1.09 | 410.68 | 0.74 |
| Oz | Intercept | -23.76 | -24.68 | -22.84 | 0.47 | 82992.47 | 31.92 | 2600.06 | 0.95 |
| Oz | Brightness | 2.05 | 1.87 | 2.23 | 0.09 | 617.64 | 1.22 | 504.68 | 0.78 |
| P3 | Intercept | -28.91 | -29.57 | -28.26 | 0.33 | 122874.54 | 16.20 | 7583.54 | 0.98 |
| P3 | Brightness | 1.08 | 0.93 | 1.22 | 0.07 | 170.46 | 0.82 | 207.32 | 0.59 |
| P4 | Intercept | -27.08 | -27.83 | -26.33 | 0.38 | 107831.97 | 21.16 | 5096.26 | 0.97 |
| P4 | Brightness | 1.18 | 1.04 | 1.32 | 0.07 | 203.70 | 0.72 | 283.55 | 0.66 |

| | | | | | | | | | |
|---|---|---|---|---|---|---|---|---|---|
| P7 | Intercept | -28.41 | -29.16 | -27.66 | 0.38 | 118621.46 | 21.12 | 5617.08 | 0.97 |
| P7 | Brightness | 1.40 | 1.25 | 1.56 | 0.08 | 289.50 | 0.87 | 331.65 | 0.69 |
| P8 | Intercept | -27.23 | -28.02 | -26.44 | 0.40 | 109028.88 | 23.47 | 4646.25 | 0.97 |
| P8 | Brightness | 1.35 | 1.20 | 1.50 | 0.08 | 269.01 | 0.84 | 321.58 | 0.69 |
| Pz | Intercept | -27.99 | -28.68 | -27.31 | 0.35 | 115184.29 | 17.76 | 6484.82 | 0.98 |
| Pz | Brightness | 0.99 | 0.83 | 1.14 | 0.08 | 143.36 | 0.89 | 160.59 | 0.52 |
| T7 | Intercept | -32.69 | -33.35 | -32.03 | 0.34 | 157068.91 | 16.51 | 9512.60 | 0.98 |
| T7 | Brightness | 0.59 | 0.48 | 0.70 | 0.06 | 50.65 | 0.45 | 111.63 | 0.43 |
| T8 | Intercept | -33.08 | -33.73 | -32.42 | 0.33 | 160826.65 | 16.17 | 9946.21 | 0.99 |
| T8 | Brightness | 0.81 | 0.68 | 0.93 | 0.06 | 95.44 | 0.62 | 155.18 | 0.52 |
| TP10 | Intercept | -30.83 | -31.54 | -30.11 | 0.36 | 139682.32 | 19.46 | 7178.68 | 0.98 |
| TP10 | Brightness | 1.26 | 1.11 | 1.42 | 0.08 | 233.98 | 0.91 | 258.47 | 0.64 |
| TP9 | Intercept | -31.11 | -31.78 | -30.43 | 0.34 | 142226.41 | 17.32 | 8213.65 | 0.98 |
| TP9 | Brightness | 0.91 | 0.77 | 1.05 | 0.07 | 122.67 | 0.75 | 164.05 | 0.53 |

*Note*. Both FDR corrected and uncorrected $p$ values were < .001; $df$ for the numerator and denominator were 1 and 46, respectively; LL = lower level; UL = upper level.

Supplementary Table 3

*Analysis 2 Electrode-wise Multilevel Modeling Results: Estimating Brain-Behavior Relationships and the Moderating Effects of Pain and Somatic Symptoms*

| | | | | 95% CI | | | | | | | | |
|---|---|---|---|---|---|---|---|---|---|---|---|---|
| Electrode | Model | Source | *b* | *LL* | *UL* | *SE* | *SS* | *MSE* | *F* | *p* | $p_{fdr}$ | $\eta_p^2$ |
| All | Intercept | Intercept | 7.997 | 7.367 | 8.627 | 0.319 | 9401.60 | 14.92 | 630.0 | < . 001 | < . 001 | .815 |
| All | Intercept | Bladder Pain | 0.064 | 0.022 | 0.107 | 0.021 | 134.33 | 14.92 | 9.0 | .003 | .003 | .059 |
| All | Intercept | Menstrual Pain | 0.012 | -0.013 | 0.038 | 0.013 | 13.99 | 14.92 | 0.9 | .335 | .335 | .007 |
| All | Intercept | Somatic Symptoms | -0.135 | -0.411 | 0.140 | 0.139 | 14.13 | 14.92 | 0.9 | .332 | .332 | .007 |
| C3 | Brightness | Intercept | 0.559 | 0.411 | 0.706 | 0.075 | 45.92 | 0.82 | 56.1 | < . 001 | < . 001 | .282 |
| C3 | Brightness | Bladder Pain | 0.000 | -0.010 | 0.010 | 0.005 | 0.00 | 0.82 | 0.0 | .985 | .985 | < . 001 |
| C3 | Brightness | Menstrual Pain | 0.005 | -0.001 | 0.011 | 0.003 | 2.35 | 0.82 | 2.9 | .093 | .153 | .02 |
| C3 | Brightness | Somatic Symptoms | -0.028 | -0.093 | 0.036 | 0.033 | 0.61 | 0.82 | 0.7 | .389 | .817 | .005 |
| C4 | Brightness | Intercept | 0.451 | 0.297 | 0.606 | 0.078 | 29.96 | 0.90 | 33.4 | < . 001 | < . 001 | .189 |
| C4 | Brightness | Bladder Pain | -0.005 | -0.015 | 0.005 | 0.005 | 0.82 | 0.90 | 0.9 | .34 | .985 | .006 |
| C4 | Brightness | Menstrual Pain | 0.006 | 0.000 | 0.012 | 0.003 | 3.40 | 0.90 | 3.8 | .054 | .132 | .026 |
| C4 | Brightness | Somatic Symptoms | -0.053 | -0.121 | 0.014 | 0.034 | 2.18 | 0.90 | 2.4 | .121 | .58 | .017 |
| CP1 | Brightness | Intercept | 0.492 | 0.351 | 0.633 | 0.071 | 35.61 | 0.75 | 47.7 | < . 001 | < . 001 | .25 |
| CP1 | Brightness | Bladder Pain | -0.002 | -0.011 | 0.008 | 0.005 | 0.10 | 0.75 | 0.1 | .717 | .985 | .001 |
| CP1 | Brightness | Menstrual Pain | 0.005 | -0.001 | 0.010 | 0.003 | 1.99 | 0.75 | 2.7 | .105 | .153 | .018 |
| CP1 | Brightness | Somatic Symptoms | 0.013 | -0.048 | 0.075 | 0.031 | 0.14 | 0.75 | 0.2 | .669 | .93 | .001 |
| CP2 | Brightness | Intercept | 0.448 | 0.256 | 0.640 | 0.097 | 29.50 | 1.38 | 21.3 | < . 001 | < . 001 | .13 |
| CP2 | Brightness | Bladder Pain | 0.001 | -0.012 | 0.014 | 0.007 | 0.01 | 1.38 | 0.0 | .926 | .985 | < . 001 |
| CP2 | Brightness | Menstrual Pain | 0.008 | 0.001 | 0.016 | 0.004 | 6.26 | 1.38 | 4.5 | .035 | .126 | .031 |
| CP2 | Brightness | Somatic Symptoms | -0.078 | -0.162 | 0.006 | 0.042 | 4.71 | 1.38 | 3.4 | .067 | .535 | .023 |
| CP5 | Brightness | Intercept | 0.434 | 0.262 | 0.606 | 0.087 | 27.64 | 1.11 | 24.8 | < . 001 | < . 001 | .148 |
| CP5 | Brightness | Bladder Pain | 0.002 | -0.010 | 0.013 | 0.006 | 0.09 | 1.11 | 0.1 | .78 | .985 | .001 |
| CP5 | Brightness | Menstrual Pain | 0.004 | -0.003 | 0.011 | 0.004 | 1.55 | 1.11 | 1.4 | .239 | .278 | .01 |

| | | | | | | | | | | | |
|---|---|---|---|---|---|---|---|---|---|---|---|
| CP5 | Brightness | Somatic Symptoms | -0.029 | -0.104 | 0.046 | 0.038 | 0.64 | 1.11 | 0.6 | .448 | .843 | .004 |
| CP6 | Brightness | Intercept | 0.428 | 0.240 | 0.617 | 0.095 | 26.96 | 1.34 | 20.2 | < . 001 | < . 001 | .124 |
| CP6 | Brightness | Bladder Pain | 0.005 | -0.007 | 0.018 | 0.006 | 0.96 | 1.34 | 0.7 | .398 | .985 | .005 |
| CP6 | Brightness | Menstrual Pain | 0.006 | -0.001 | 0.014 | 0.004 | 3.71 | 1.34 | 2.8 | .098 | .153 | .019 |
| CP6 | Brightness | Somatic Symptoms | -0.157 | -0.239 | -0.074 | 0.042 | 18.87 | 1.34 | 14.1 | < . 001 | .008 | .09 |
| Cz | Brightness | Intercept | 0.481 | 0.333 | 0.629 | 0.075 | 33.98 | 0.82 | 41.2 | < . 001 | < . 001 | .224 |
| Cz | Brightness | Bladder Pain | -0.002 | -0.012 | 0.008 | 0.005 | 0.09 | 0.82 | 0.1 | .747 | .985 | .001 |
| Cz | Brightness | Menstrual Pain | 0.004 | -0.002 | 0.010 | 0.003 | 1.63 | 0.82 | 2.0 | .162 | .215 | .014 |
| Cz | Brightness | Somatic Symptoms | -0.018 | -0.083 | 0.046 | 0.033 | 0.25 | 0.82 | 0.3 | .579 | .896 | .002 |
| F3 | Brightness | Intercept | 0.562 | 0.423 | 0.700 | 0.070 | 46.37 | 0.72 | 64.2 | < . 001 | < . 001 | .31 |
| F3 | Brightness | Bladder Pain | -0.001 | -0.010 | 0.009 | 0.005 | 0.01 | 0.72 | 0.0 | .913 | .985 | < . 001 |
| F3 | Brightness | Menstrual Pain | 0.009 | 0.003 | 0.014 | 0.003 | 6.70 | 0.72 | 9.3 | .003 | .089 | .061 |
| F3 | Brightness | Somatic Symptoms | -0.008 | -0.068 | 0.053 | 0.031 | 0.04 | 0.72 | 0.1 | .804 | .934 | < . 001 |
| F4 | Brightness | Intercept | 0.485 | 0.357 | 0.613 | 0.065 | 34.58 | 0.62 | 56.1 | < . 001 | < . 001 | .282 |
| F4 | Brightness | Bladder Pain | 0.002 | -0.006 | 0.011 | 0.004 | 0.16 | 0.62 | 0.3 | .61 | .985 | .002 |
| F4 | Brightness | Menstrual Pain | 0.005 | 0.000 | 0.010 | 0.003 | 2.15 | 0.62 | 3.5 | .064 | .134 | .024 |
| F4 | Brightness | Somatic Symptoms | -0.032 | -0.088 | 0.024 | 0.028 | 0.79 | 0.62 | 1.3 | .259 | .723 | .009 |
| F7 | Brightness | Intercept | 0.515 | 0.363 | 0.667 | 0.077 | 39.00 | 0.87 | 44.7 | < . 001 | < . 001 | .238 |
| F7 | Brightness | Bladder Pain | 0.007 | -0.004 | 0.017 | 0.005 | 1.43 | 0.87 | 1.6 | .203 | .985 | .011 |
| F7 | Brightness | Menstrual Pain | 0.004 | -0.002 | 0.010 | 0.003 | 1.40 | 0.87 | 1.6 | .208 | .266 | .011 |
| F7 | Brightness | Somatic Symptoms | -0.016 | -0.083 | 0.051 | 0.034 | 0.20 | 0.87 | 0.2 | .635 | .924 | .002 |
| F8 | Brightness | Intercept | 0.450 | 0.309 | 0.590 | 0.071 | 29.75 | 0.74 | 40.0 | < . 001 | < . 001 | .219 |
| F8 | Brightness | Bladder Pain | 0.003 | -0.006 | 0.013 | 0.005 | 0.37 | 0.74 | 0.5 | .484 | .985 | .003 |
| F8 | Brightness | Menstrual Pain | 0.002 | -0.003 | 0.008 | 0.003 | 0.55 | 0.74 | 0.7 | .392 | .433 | .005 |
| F8 | Brightness | Somatic Symptoms | 0.006 | -0.056 | 0.067 | 0.031 | 0.03 | 0.74 | 0.0 | .85 | .934 | < . 001 |
| FC1 | Brightness | Intercept | 0.685 | 0.490 | 0.879 | 0.098 | 68.91 | 1.42 | 48.5 | < . 001 | < . 001 | .253 |
| FC1 | Brightness | Bladder Pain | 0.007 | -0.006 | 0.020 | 0.007 | 1.62 | 1.42 | 1.1 | .288 | .985 | .008 |
| FC1 | Brightness | Menstrual Pain | 0.008 | 0.001 | 0.016 | 0.004 | 6.40 | 1.42 | 4.5 | .036 | .126 | .031 |
| FC1 | Brightness | Somatic Symptoms | -0.006 | -0.091 | 0.079 | 0.043 | 0.03 | 1.42 | 0.0 | .886 | .934 | < . 001 |
| FC2 | Brightness | Intercept | 0.526 | 0.386 | 0.666 | 0.071 | 40.72 | 0.74 | 55.2 | < . 001 | < . 001 | .278 |
| FC2 | Brightness | Bladder Pain | 0.003 | -0.007 | 0.012 | 0.005 | 0.22 | 0.74 | 0.3 | .585 | .985 | .002 |

| | | | | | | | | | | | | |
|---|---|---|---|---|---|---|---|---|---|---|---|---|
| FC2 | Brightness | Menstrual Pain | 0.003 | -0.002 | 0.009 | 0.003 | 1.08 | 0.74 | 1.5 | .228 | .278 | .01 |
| FC2 | Brightness | Somatic Symptoms | -0.010 | -0.071 | 0.052 | 0.031 | 0.07 | 0.74 | 0.1 | .756 | .934 | .001 |
| FC5 | Brightness | Intercept | 0.479 | 0.349 | 0.608 | 0.065 | 33.69 | 0.63 | 53.6 | < . 001 | < . 001 | .273 |
| FC5 | Brightness | Bladder Pain | 0.002 | -0.007 | 0.011 | 0.004 | 0.13 | 0.63 | 0.2 | .654 | .985 | .001 |
| FC5 | Brightness | Menstrual Pain | 0.004 | -0.001 | 0.010 | 0.003 | 1.68 | 0.63 | 2.7 | .104 | .153 | .018 |
| FC5 | Brightness | Somatic Symptoms | -0.018 | -0.075 | 0.038 | 0.029 | 0.26 | 0.63 | 0.4 | .519 | .896 | .003 |
| FC6 | Brightness | Intercept | 0.527 | 0.394 | 0.660 | 0.067 | 40.86 | 0.67 | 61.4 | < . 001 | < . 001 | .3 |
| FC6 | Brightness | Bladder Pain | 0.001 | -0.008 | 0.010 | 0.005 | 0.05 | 0.67 | 0.1 | .789 | .985 | .001 |
| FC6 | Brightness | Menstrual Pain | 0.002 | -0.004 | 0.007 | 0.003 | 0.22 | 0.67 | 0.3 | .565 | .583 | .002 |
| FC6 | Brightness | Somatic Symptoms | -0.016 | -0.074 | 0.042 | 0.029 | 0.20 | 0.67 | 0.3 | .588 | .896 | .002 |
| Fp1 | Brightness | Intercept | 0.524 | 0.361 | 0.688 | 0.083 | 40.43 | 1.01 | 40.1 | < . 001 | < . 001 | .219 |
| Fp1 | Brightness | Bladder Pain | 0.001 | -0.010 | 0.012 | 0.006 | 0.03 | 1.01 | 0.0 | .863 | .985 | < . 001 |
| Fp1 | Brightness | Menstrual Pain | 0.008 | 0.001 | 0.015 | 0.003 | 5.82 | 1.01 | 5.8 | .018 | .113 | .039 |
| Fp1 | Brightness | Somatic Symptoms | -0.072 | -0.144 | -0.001 | 0.036 | 4.01 | 1.01 | 4.0 | .048 | .514 | .027 |
| Fp2 | Brightness | Intercept | 0.428 | 0.279 | 0.578 | 0.076 | 26.98 | 0.84 | 32.0 | < . 001 | < . 001 | .183 |
| Fp2 | Brightness | Bladder Pain | 0.004 | -0.006 | 0.014 | 0.005 | 0.44 | 0.84 | 0.5 | .474 | .985 | .004 |
| Fp2 | Brightness | Menstrual Pain | 0.004 | -0.002 | 0.010 | 0.003 | 1.76 | 0.84 | 2.1 | .151 | .211 | .014 |
| Fp2 | Brightness | Somatic Symptoms | -0.011 | -0.077 | 0.054 | 0.033 | 0.10 | 0.84 | 0.1 | .732 | .934 | .001 |
| FT10 | Brightness | Intercept | 0.451 | 0.302 | 0.600 | 0.076 | 29.93 | 0.84 | 35.7 | < . 001 | < . 001 | .2 |
| FT10 | Brightness | Bladder Pain | 0.005 | -0.005 | 0.015 | 0.005 | 0.87 | 0.84 | 1.0 | .31 | .985 | .007 |
| FT10 | Brightness | Menstrual Pain | 0.005 | -0.001 | 0.011 | 0.003 | 2.29 | 0.84 | 2.7 | .1 | .153 | .019 |
| FT10 | Brightness | Somatic Symptoms | 0.002 | -0.063 | 0.067 | 0.033 | 0.00 | 0.84 | 0.0 | .955 | .955 | < . 001 |
| FT9 | Brightness | Intercept | 0.542 | 0.402 | 0.682 | 0.071 | 43.16 | 0.74 | 58.3 | < . 001 | < . 001 | .289 |
| FT9 | Brightness | Bladder Pain | 0.003 | -0.006 | 0.013 | 0.005 | 0.32 | 0.74 | 0.4 | .512 | .985 | .003 |
| FT9 | Brightness | Menstrual Pain | 0.006 | 0.000 | 0.012 | 0.003 | 3.20 | 0.74 | 4.3 | .039 | .126 | .029 |
| FT9 | Brightness | Somatic Symptoms | -0.046 | -0.107 | 0.015 | 0.031 | 1.64 | 0.74 | 2.2 | .139 | .58 | .015 |
| Fz | Brightness | Intercept | 0.515 | 0.373 | 0.657 | 0.072 | 39.02 | 0.76 | 51.5 | < . 001 | < . 001 | .265 |
| Fz | Brightness | Bladder Pain | 0.005 | -0.005 | 0.014 | 0.005 | 0.67 | 0.76 | 0.9 | .35 | .985 | .006 |
| Fz | Brightness | Menstrual Pain | 0.006 | 0.000 | 0.011 | 0.003 | 2.90 | 0.76 | 3.8 | .052 | .132 | .026 |
| Fz | Brightness | Somatic Symptoms | -0.029 | -0.091 | 0.033 | 0.031 | 0.67 | 0.76 | 0.9 | .35 | .817 | .006 |
| O1 | Brightness | Intercept | 0.459 | 0.301 | 0.618 | 0.080 | 31.03 | 0.95 | 32.7 | < . 001 | < . 001 | .186 |

| | | | | | | | | | | | |
|---|---|---|---|---|---|---|---|---|---|---|---|
| O1 | Brightness | Bladder Pain | -0.001 | -0.011 | 0.010 | 0.005 | 0.01 | 0.95 | 0.0 | .905 | .985 | . |
| O1 | Brightness | Menstrual Pain | 0.008 | 0.002 | 0.015 | 0.003 | 6.42 | 0.95 | 6.8 | .01 | .109 | .045 |
| O1 | Brightness | Somatic Symptoms | -0.059 | -0.129 | 0.010 | 0.035 | 2.69 | 0.95 | 2.8 | .094 | .58 | .019 |
| O2 | Brightness | Intercept | 0.410 | 0.258 | 0.562 | 0.077 | 24.69 | 0.87 | 28.4 | < . 001 | < . 001 | .166 |
| O2 | Brightness | Bladder Pain | 0.005 | -0.005 | 0.016 | 0.005 | 0.94 | 0.87 | 1.1 | .3 | .985 | .008 |
| O2 | Brightness | Menstrual Pain | 0.004 | -0.002 | 0.010 | 0.003 | 1.19 | 0.87 | 1.4 | .243 | .278 | .01 |
| O2 | Brightness | Somatic Symptoms | -0.031 | -0.097 | 0.036 | 0.034 | 0.74 | 0.87 | 0.8 | .359 | .817 | .006 |
| Oz | Brightness | Intercept | 0.357 | 0.225 | 0.489 | 0.067 | 18.75 | 0.66 | 28.6 | < . 001 | < . 001 | .167 |
| Oz | Brightness | Bladder Pain | -0.003 | -0.012 | 0.006 | 0.004 | 0.27 | 0.66 | 0.4 | .519 | .985 | .003 |
| Oz | Brightness | Menstrual Pain | 0.006 | 0.001 | 0.011 | 0.003 | 3.32 | 0.66 | 5.1 | .026 | .118 | .034 |
| Oz | Brightness | Somatic Symptoms | -0.032 | -0.090 | 0.025 | 0.029 | 0.80 | 0.66 | 1.2 | .271 | .723 | .008 |
| P3 | Brightness | Intercept | 0.453 | 0.305 | 0.601 | 0.075 | 30.18 | 0.82 | 36.7 | < . 001 | < . 001 | .204 |
| P3 | Brightness | Bladder Pain | 0.000 | -0.010 | 0.010 | 0.005 | 0.01 | 0.82 | 0.0 | .933 | .985 | < . 001 |
| P3 | Brightness | Menstrual Pain | 0.006 | 0.000 | 0.012 | 0.003 | 3.22 | 0.82 | 3.9 | .05 | .132 | .027 |
| P3 | Brightness | Somatic Symptoms | -0.042 | -0.106 | 0.023 | 0.033 | 1.35 | 0.82 | 1.6 | .202 | .718 | .011 |
| P4 | Brightness | Intercept | 0.481 | 0.345 | 0.616 | 0.069 | 34.01 | 0.69 | 49.2 | < . 001 | < . 001 | .256 |
| P4 | Brightness | Bladder Pain | 0.006 | -0.003 | 0.015 | 0.005 | 1.24 | 0.69 | 1.8 | .182 | .985 | .012 |
| P4 | Brightness | Menstrual Pain | 0.006 | 0.001 | 0.012 | 0.003 | 3.71 | 0.69 | 5.4 | .022 | .117 | .036 |
| P4 | Brightness | Somatic Symptoms | -0.044 | -0.103 | 0.015 | 0.030 | 1.48 | 0.69 | 2.1 | .145 | .58 | .015 |
| P7 | Brightness | Intercept | 0.425 | 0.279 | 0.571 | 0.074 | 26.51 | 0.80 | 33.0 | < . 001 | < . 001 | .188 |
| P7 | Brightness | Bladder Pain | -0.003 | -0.013 | 0.007 | 0.005 | 0.28 | 0.80 | 0.3 | .559 | .985 | .002 |
| P7 | Brightness | Menstrual Pain | 0.008 | 0.002 | 0.014 | 0.003 | 6.34 | 0.80 | 7.9 | .006 | .09 | .052 |
| P7 | Brightness | Somatic Symptoms | -0.027 | -0.091 | 0.037 | 0.032 | 0.55 | 0.80 | 0.7 | .408 | .817 | .005 |
| P8 | Brightness | Intercept | 0.437 | 0.298 | 0.576 | 0.070 | 28.11 | 0.73 | 38.6 | < . 001 | < . 001 | .213 |
| P8 | Brightness | Bladder Pain | 0.003 | -0.006 | 0.012 | 0.005 | 0.29 | 0.73 | 0.4 | .525 | .985 | .003 |
| P8 | Brightness | Menstrual Pain | 0.007 | 0.001 | 0.012 | 0.003 | 4.20 | 0.73 | 5.8 | .018 | .113 | .039 |
| P8 | Brightness | Somatic Symptoms | -0.078 | -0.139 | -0.017 | 0.031 | 4.68 | 0.73 | 6.4 | .012 | .197 | .043 |
| Pz | Brightness | Intercept | 0.425 | 0.291 | 0.560 | 0.068 | 26.56 | 0.68 | 39.1 | < . 001 | < . 001 | .215 |
| Pz | Brightness | Bladder Pain | 0.000 | -0.009 | 0.009 | 0.005 | 0.00 | 0.68 | 0.0 | .964 | .985 | < . 001 |
| Pz | Brightness | Menstrual Pain | 0.005 | 0.000 | 0.010 | 0.003 | 2.32 | 0.68 | 3.4 | .067 | .134 | .023 |
| Pz | Brightness | Somatic Symptoms | -0.017 | -0.076 | 0.042 | 0.030 | 0.22 | 0.68 | 0.3 | .568 | .896 | .002 |

| | | | | | | | | | | | | |
|---|---|---|---|---|---|---|---|---|---|---|---|---|
| T7 | Brightness | Intercept | 0.341 | 0.147 | 0.535 | 0.098 | 17.09 | 1.41 | 12.1 | .001 | .001 | .078 |
| T7 | Brightness | Bladder Pain | 0.000 | -0.013 | 0.013 | 0.007 | 0.00 | 1.41 | 0.0 | .979 | .985 | < . 001 |
| T7 | Brightness | Menstrual Pain | 0.001 | -0.007 | 0.009 | 0.004 | 0.12 | 1.41 | 0.1 | .773 | .773 | .001 |
| T7 | Brightness | Somatic Symptoms | -0.007 | -0.091 | 0.078 | 0.043 | 0.03 | 1.41 | 0.0 | .877 | .934 | < . 001 |
| T8 | Brightness | Intercept | 0.459 | 0.312 | 0.605 | 0.074 | 30.94 | 0.81 | 38.4 | < . 001 | < . 001 | .211 |
| T8 | Brightness | Bladder Pain | 0.004 | -0.006 | 0.013 | 0.005 | 0.41 | 0.81 | 0.5 | .478 | .985 | .004 |
| T8 | Brightness | Menstrual Pain | 0.006 | 0.000 | 0.011 | 0.003 | 2.75 | 0.81 | 3.4 | .067 | .134 | .023 |
| T8 | Brightness | Somatic Symptoms | 0.009 | -0.055 | 0.073 | 0.032 | 0.06 | 0.81 | 0.1 | .779 | .934 | .001 |
| TP10 | Brightness | Intercept | 0.410 | 0.261 | 0.558 | 0.075 | 24.67 | 0.83 | 29.7 | < . 001 | < . 001 | .172 |
| TP10 | Brightness | Bladder Pain | 0.004 | -0.006 | 0.014 | 0.005 | 0.53 | 0.83 | 0.6 | .427 | .985 | .004 |
| TP10 | Brightness | Menstrual Pain | 0.002 | -0.004 | 0.008 | 0.003 | 0.40 | 0.83 | 0.5 | .49 | .523 | .003 |
| TP10 | Brightness | Somatic Symptoms | 0.004 | -0.061 | 0.069 | 0.033 | 0.01 | 0.83 | 0.0 | .905 | .934 | < . 001 |
| TP9 | Brightness | Intercept | 0.617 | 0.375 | 0.860 | 0.123 | 56.05 | 2.22 | 25.3 | < . 001 | < . 001 | .15 |
| TP9 | Brightness | Bladder Pain | 0.007 | -0.009 | 0.023 | 0.008 | 1.59 | 2.22 | 0.7 | .398 | .985 | .005 |
| TP9 | Brightness | Menstrual Pain | 0.008 | -0.002 | 0.018 | 0.005 | 6.17 | 2.22 | 2.8 | .097 | .153 | .019 |
| TP9 | Brightness | Somatic Symptoms | -0.061 | -0.167 | 0.045 | 0.054 | 2.85 | 2.22 | 1.3 | .258 | .723 | .009 |
| C3 | PSD | Intercept | 0.010 | -0.156 | 0.175 | 0.084 | 0.01 | 1.03 | 0.0 | .91 | .945 | < . 001 |
| C3 | PSD | Bladder Pain | 0.001 | -0.010 | 0.012 | 0.006 | 0.05 | 1.03 | 0.1 | .819 | .974 | < . 001 |
| C3 | PSD | Menstrual Pain | 0.000 | -0.007 | 0.006 | 0.003 | 0.02 | 1.03 | 0.0 | .89 | .968 | < . 001 |
| C3 | PSD | Somatic Symptoms | -0.023 | -0.096 | 0.049 | 0.037 | 0.42 | 1.03 | 0.4 | .522 | .93 | .003 |
| C4 | PSD | Intercept | 0.225 | 0.079 | 0.370 | 0.074 | 7.42 | 0.80 | 9.3 | .003 | .021 | .061 |
| C4 | PSD | Bladder Pain | 0.009 | -0.001 | 0.019 | 0.005 | 2.54 | 0.80 | 3.2 | .076 | .796 | .022 |
| C4 | PSD | Menstrual Pain | 0.002 | -0.004 | 0.008 | 0.003 | 0.34 | 0.80 | 0.4 | .516 | .968 | .003 |
| C4 | PSD | Somatic Symptoms | -0.022 | -0.086 | 0.041 | 0.032 | 0.38 | 0.80 | 0.5 | .491 | .93 | .003 |
| CP1 | PSD | Intercept | 0.041 | -0.166 | 0.248 | 0.105 | 0.25 | 1.62 | 0.2 | .697 | .779 | .001 |
| CP1 | PSD | Bladder Pain | -0.011 | -0.025 | 0.003 | 0.007 | 3.63 | 1.62 | 2.2 | .137 | .796 | .015 |
| CP1 | PSD | Menstrual Pain | 0.003 | -0.005 | 0.012 | 0.004 | 1.09 | 1.62 | 0.7 | .412 | .968 | .005 |
| CP1 | PSD | Somatic Symptoms | -0.140 | -0.231 | -0.050 | 0.046 | 15.13 | 1.62 | 9.4 | .003 | .043 | .061 |
| CP2 | PSD | Intercept | 0.047 | -0.117 | 0.211 | 0.083 | 0.33 | 1.01 | 0.3 | .569 | .701 | .002 |
| CP2 | PSD | Bladder Pain | 0.005 | -0.006 | 0.016 | 0.006 | 0.72 | 1.01 | 0.7 | .4 | .916 | .005 |
| CP2 | PSD | Menstrual Pain | -0.005 | -0.012 | 0.001 | 0.003 | 2.73 | 1.01 | 2.7 | .103 | .968 | .019 |

| | | | | | | | | | | | |
|---|---|---|---|---|---|---|---|---|---|---|---|
| CP2 | PSD | Somatic Symptoms | 0.051 | -0.021 | 0.122 | 0.036 | 1.97 | 1.01 | 1.9 | .165 | .794 | .013 |
| CP5 | PSD | Intercept | 0.125 | -0.024 | 0.274 | 0.076 | 2.29 | 0.84 | 2.7 | .1 | .268 | .019 |
| CP5 | PSD | Bladder Pain | 0.001 | -0.009 | 0.011 | 0.005 | 0.01 | 0.84 | 0.0 | .909 | .974 | < . 001 |
| CP5 | PSD | Menstrual Pain | 0.001 | -0.005 | 0.007 | 0.003 | 0.03 | 0.84 | 0.0 | .858 | .968 | < . 001 |
| CP5 | PSD | Somatic Symptoms | -0.035 | -0.100 | 0.031 | 0.033 | 0.92 | 0.84 | 1.1 | .297 | .794 | .008 |
| CP6 | PSD | Intercept | 0.202 | 0.091 | 0.314 | 0.056 | 6.01 | 0.47 | 12.9 | < . 001 | .012 | .083 |
| CP6 | PSD | Bladder Pain | -0.001 | -0.009 | 0.006 | 0.004 | 0.05 | 0.47 | 0.1 | .741 | .974 | .001 |
| CP6 | PSD | Menstrual Pain | 0.000 | -0.005 | 0.004 | 0.002 | 0.00 | 0.47 | 0.0 | .968 | .968 | < . 001 |
| CP6 | PSD | Somatic Symptoms | 0.088 | 0.040 | 0.137 | 0.025 | 5.99 | 0.47 | 12.8 | < . 001 | .015 | .082 |
| Cz | PSD | Intercept | 0.062 | -0.091 | 0.215 | 0.077 | 0.57 | 0.88 | 0.6 | .423 | .618 | .005 |
| Cz | PSD | Bladder Pain | 0.006 | -0.004 | 0.017 | 0.005 | 1.31 | 0.88 | 1.5 | .225 | .796 | .01 |
| Cz | PSD | Menstrual Pain | 0.000 | -0.006 | 0.006 | 0.003 | 0.00 | 0.88 | 0.0 | .948 | .968 | < . 001 |
| Cz | PSD | Somatic Symptoms | 0.009 | -0.058 | 0.076 | 0.034 | 0.06 | 0.88 | 0.1 | .79 | .931 | < . 001 |
| F3 | PSD | Intercept | 0.039 | -0.146 | 0.224 | 0.094 | 0.22 | 1.29 | 0.2 | .68 | .779 | .001 |
| F3 | PSD | Bladder Pain | 0.001 | -0.011 | 0.014 | 0.006 | 0.05 | 1.29 | 0.0 | .849 | .974 | < . 001 |
| F3 | PSD | Menstrual Pain | 0.001 | -0.007 | 0.008 | 0.004 | 0.03 | 1.29 | 0.0 | .88 | .968 | < . 001 |
| F3 | PSD | Somatic Symptoms | -0.053 | -0.134 | 0.028 | 0.041 | 2.15 | 1.29 | 1.7 | .199 | .794 | .012 |
| F4 | PSD | Intercept | 0.176 | -0.024 | 0.376 | 0.101 | 4.57 | 1.50 | 3.0 | .083 | .242 | .021 |
| F4 | PSD | Bladder Pain | -0.001 | -0.015 | 0.012 | 0.007 | 0.05 | 1.50 | 0.0 | .856 | .974 | < . 001 |
| F4 | PSD | Menstrual Pain | -0.002 | -0.010 | 0.006 | 0.004 | 0.43 | 1.50 | 0.3 | .594 | .968 | .002 |
| F4 | PSD | Somatic Symptoms | -0.017 | -0.104 | 0.070 | 0.044 | 0.22 | 1.50 | 0.1 | .7 | .931 | .001 |
| F7 | PSD | Intercept | 0.150 | -0.044 | 0.343 | 0.098 | 3.29 | 1.41 | 2.3 | .129 | .283 | .016 |
| F7 | PSD | Bladder Pain | 0.003 | -0.010 | 0.016 | 0.007 | 0.22 | 1.41 | 0.2 | .692 | .974 | .001 |
| F7 | PSD | Menstrual Pain | -0.001 | -0.008 | 0.007 | 0.004 | 0.03 | 1.41 | 0.0 | .893 | .968 | < . 001 |
| F7 | PSD | Somatic Symptoms | -0.003 | -0.088 | 0.081 | 0.043 | 0.01 | 1.41 | 0.0 | .941 | .941 | < . 001 |
| F8 | PSD | Intercept | 0.195 | 0.019 | 0.371 | 0.089 | 5.61 | 1.17 | 4.8 | .03 | .16 | .033 |
| F8 | PSD | Bladder Pain | 0.002 | -0.010 | 0.014 | 0.006 | 0.10 | 1.17 | 0.1 | .767 | .974 | .001 |
| F8 | PSD | Menstrual Pain | 0.002 | -0.005 | 0.009 | 0.004 | 0.25 | 1.17 | 0.2 | .642 | .968 | .002 |
| F8 | PSD | Somatic Symptoms | -0.042 | -0.119 | 0.035 | 0.039 | 1.34 | 1.17 | 1.2 | .285 | .794 | .008 |
| FC1 | PSD | Intercept | -0.210 | -0.594 | 0.173 | 0.194 | 6.50 | 5.54 | 1.2 | .28 | .472 | .008 |
| FC1 | PSD | Bladder Pain | -0.017 | -0.043 | 0.008 | 0.013 | 9.89 | 5.54 | 1.8 | .184 | .796 | .012 |

| | | | | | | | | | | | | |
|---|---|---|---|---|---|---|---|---|---|---|---|---|
| FC1 | PSD | Menstrual Pain | -0.007 | -0.022 | 0.009 | 0.008 | 4.08 | 5.54 | 0.7 | .392 | .968 | .005 |
| FC1 | PSD | Somatic Symptoms | 0.042 | -0.126 | 0.210 | 0.085 | 1.36 | 5.54 | 0.2 | .62 | .931 | .002 |
| FC2 | PSD | Intercept | 0.119 | -0.070 | 0.308 | 0.096 | 2.09 | 1.35 | 1.6 | .215 | .404 | .011 |
| FC2 | PSD | Bladder Pain | -0.007 | -0.020 | 0.005 | 0.006 | 1.80 | 1.35 | 1.3 | .249 | .796 | .009 |
| FC2 | PSD | Menstrual Pain | 0.004 | -0.003 | 0.012 | 0.004 | 1.66 | 1.35 | 1.2 | .268 | .968 | .009 |
| FC2 | PSD | Somatic Symptoms | -0.035 | -0.117 | 0.048 | 0.042 | 0.93 | 1.35 | 0.7 | .407 | .868 | .005 |
| FC5 | PSD | Intercept | -0.006 | -0.172 | 0.160 | 0.084 | 0.00 | 1.04 | 0.0 | .945 | .945 | < . 001 |
| FC5 | PSD | Bladder Pain | -0.007 | -0.018 | 0.005 | 0.006 | 1.44 | 1.04 | 1.4 | .241 | .796 | .01 |
| FC5 | PSD | Menstrual Pain | 0.004 | -0.002 | 0.011 | 0.003 | 1.80 | 1.04 | 1.7 | .189 | .968 | .012 |
| FC5 | PSD | Somatic Symptoms | -0.010 | -0.083 | 0.062 | 0.037 | 0.08 | 1.04 | 0.1 | .782 | .931 | .001 |
| FC6 | PSD | Intercept | 0.008 | -0.162 | 0.179 | 0.086 | 0.01 | 1.09 | 0.0 | .924 | .945 | < . 001 |
| FC6 | PSD | Bladder Pain | 0.010 | -0.001 | 0.022 | 0.006 | 3.51 | 1.09 | 3.2 | .075 | .796 | .022 |
| FC6 | PSD | Menstrual Pain | 0.001 | -0.006 | 0.008 | 0.003 | 0.15 | 1.09 | 0.1 | .716 | .968 | .001 |
| FC6 | PSD | Somatic Symptoms | 0.007 | -0.068 | 0.082 | 0.038 | 0.04 | 1.09 | 0.0 | .853 | .931 | < . 001 |
| Fp1 | PSD | Intercept | 0.035 | -0.150 | 0.221 | 0.094 | 0.18 | 1.29 | 0.1 | .706 | .779 | .001 |
| Fp1 | PSD | Bladder Pain | 0.006 | -0.007 | 0.018 | 0.006 | 1.00 | 1.29 | 0.8 | .381 | .916 | .005 |
| Fp1 | PSD | Menstrual Pain | -0.001 | -0.008 | 0.007 | 0.004 | 0.03 | 1.29 | 0.0 | .887 | .968 | < . 001 |
| Fp1 | PSD | Somatic Symptoms | 0.054 | -0.027 | 0.135 | 0.041 | 2.26 | 1.29 | 1.8 | .188 | .794 | .012 |
| Fp2 | PSD | Intercept | 0.092 | -0.141 | 0.325 | 0.118 | 1.25 | 2.04 | 0.6 | .435 | .618 | .004 |
| Fp2 | PSD | Bladder Pain | 0.004 | -0.011 | 0.020 | 0.008 | 0.59 | 2.04 | 0.3 | .593 | .974 | .002 |
| Fp2 | PSD | Menstrual Pain | 0.001 | -0.009 | 0.010 | 0.005 | 0.05 | 2.04 | 0.0 | .874 | .968 | < . 001 |
| Fp2 | PSD | Somatic Symptoms | -0.060 | -0.161 | 0.042 | 0.052 | 2.74 | 2.04 | 1.3 | .249 | .794 | .009 |
| FT10 | PSD | Intercept | 0.152 | -0.010 | 0.315 | 0.082 | 3.42 | 0.99 | 3.5 | .065 | .236 | .024 |
| FT10 | PSD | Bladder Pain | -0.005 | -0.016 | 0.006 | 0.006 | 0.83 | 0.99 | 0.8 | .361 | .916 | .006 |
| FT10 | PSD | Menstrual Pain | 0.001 | -0.006 | 0.007 | 0.003 | 0.04 | 0.99 | 0.0 | .833 | .968 | < . 001 |
| FT10 | PSD | Somatic Symptoms | -0.037 | -0.108 | 0.034 | 0.036 | 1.07 | 0.99 | 1.1 | .299 | .794 | .008 |
| FT9 | PSD | Intercept | 0.067 | -0.146 | 0.279 | 0.108 | 0.65 | 1.70 | 0.4 | .536 | .701 | .003 |
| FT9 | PSD | Bladder Pain | 0.003 | -0.012 | 0.017 | 0.007 | 0.24 | 1.70 | 0.1 | .706 | .974 | .001 |
| FT9 | PSD | Menstrual Pain | -0.002 | -0.010 | 0.007 | 0.004 | 0.34 | 1.70 | 0.2 | .657 | .968 | .001 |
| FT9 | PSD | Somatic Symptoms | 0.080 | -0.013 | 0.172 | 0.047 | 4.87 | 1.70 | 2.9 | .093 | .794 | .02 |
| Fz | PSD | Intercept | 0.086 | -0.128 | 0.300 | 0.108 | 1.08 | 1.72 | 0.6 | .428 | .618 | .004 |

| | | | | | | | | | | | | |
|---|---|---|---|---|---|---|---|---|---|---|---|---|
| Fz | PSD | Bladder Pain | -0.005 | -0.019 | 0.009 | 0.007 | 0.84 | 1.72 | 0.5 | .486 | .973 | .003 |
| Fz | PSD | Menstrual Pain | -0.001 | -0.009 | 0.008 | 0.004 | 0.06 | 1.72 | 0.0 | .852 | .968 | < . 001 |
| Fz | PSD | Somatic Symptoms | -0.006 | -0.099 | 0.088 | 0.047 | 0.03 | 1.72 | 0.0 | .902 | .931 | < . 001 |
| O1 | PSD | Intercept | 0.089 | -0.045 | 0.222 | 0.067 | 1.15 | 0.67 | 1.7 | .192 | .383 | .012 |
| O1 | PSD | Bladder Pain | 0.007 | -0.002 | 0.016 | 0.005 | 1.50 | 0.67 | 2.2 | .137 | .796 | .015 |
| O1 | PSD | Menstrual Pain | -0.005 | -0.010 | 0.001 | 0.003 | 2.05 | 0.67 | 3.1 | .082 | .968 | .021 |
| O1 | PSD | Somatic Symptoms | 0.006 | -0.052 | 0.064 | 0.029 | 0.03 | 0.67 | 0.0 | .838 | .931 | < . 001 |
| O2 | PSD | Intercept | 0.075 | -0.057 | 0.207 | 0.067 | 0.83 | 0.65 | 1.3 | .262 | .466 | .009 |
| O2 | PSD | Bladder Pain | -0.006 | -0.014 | 0.003 | 0.004 | 0.99 | 0.65 | 1.5 | .22 | .796 | .01 |
| O2 | PSD | Menstrual Pain | 0.001 | -0.004 | 0.007 | 0.003 | 0.14 | 0.65 | 0.2 | .643 | .968 | .002 |
| O2 | PSD | Somatic Symptoms | -0.006 | -0.064 | 0.051 | 0.029 | 0.03 | 0.65 | 0.0 | .827 | .931 | < . 001 |
| Oz | PSD | Intercept | 0.107 | 0.040 | 0.174 | 0.034 | 1.68 | 0.17 | 9.9 | .002 | .021 | .065 |
| Oz | PSD | Bladder Pain | 0.005 | 0.001 | 0.010 | 0.002 | 0.83 | 0.17 | 4.9 | .029 | .796 | .033 |
| Oz | PSD | Menstrual Pain | -0.001 | -0.004 | 0.002 | 0.001 | 0.12 | 0.17 | 0.7 | .404 | .968 | .005 |
| Oz | PSD | Somatic Symptoms | -0.005 | -0.034 | 0.025 | 0.015 | 0.02 | 0.17 | 0.1 | .75 | .931 | .001 |
| P3 | PSD | Intercept | 0.100 | -0.012 | 0.212 | 0.057 | 1.46 | 0.47 | 3.1 | .081 | .242 | .021 |
| P3 | PSD | Bladder Pain | 0.001 | -0.007 | 0.008 | 0.004 | 0.02 | 0.47 | 0.1 | .821 | .974 | < . 001 |
| P3 | PSD | Menstrual Pain | 0.003 | -0.001 | 0.008 | 0.002 | 0.85 | 0.47 | 1.8 | .182 | .968 | .012 |
| P3 | PSD | Somatic Symptoms | -0.028 | -0.077 | 0.021 | 0.025 | 0.61 | 0.47 | 1.3 | .26 | .794 | .009 |
| P4 | PSD | Intercept | 0.083 | -0.025 | 0.192 | 0.055 | 1.01 | 0.44 | 2.3 | .132 | .283 | .016 |
| P4 | PSD | Bladder Pain | -0.002 | -0.009 | 0.005 | 0.004 | 0.11 | 0.44 | 0.3 | .615 | .974 | .002 |
| P4 | PSD | Menstrual Pain | -0.003 | -0.007 | 0.002 | 0.002 | 0.64 | 0.44 | 1.4 | .231 | .968 | .01 |
| P4 | PSD | Somatic Symptoms | 0.007 | -0.041 | 0.054 | 0.024 | 0.03 | 0.44 | 0.1 | .782 | .931 | .001 |
| P7 | PSD | Intercept | 0.117 | -0.001 | 0.235 | 0.060 | 2.02 | 0.52 | 3.9 | .051 | .235 | .026 |
| P7 | PSD | Bladder Pain | 0.003 | -0.005 | 0.011 | 0.004 | 0.37 | 0.52 | 0.7 | .401 | .916 | .005 |
| P7 | PSD | Menstrual Pain | -0.003 | -0.008 | 0.002 | 0.002 | 0.83 | 0.52 | 1.6 | .208 | .968 | .011 |
| P7 | PSD | Somatic Symptoms | 0.026 | -0.026 | 0.077 | 0.026 | 0.51 | 0.52 | 1.0 | .323 | .794 | .007 |
| P8 | PSD | Intercept | 0.199 | 0.085 | 0.313 | 0.058 | 5.84 | 0.49 | 11.9 | .001 | .012 | .077 |
| P8 | PSD | Bladder Pain | 0.000 | -0.008 | 0.008 | 0.004 | 0.00 | 0.49 | 0.0 | .974 | .974 | < . 001 |
| P8 | PSD | Menstrual Pain | 0.001 | -0.004 | 0.005 | 0.002 | 0.03 | 0.49 | 0.1 | .814 | .968 | < . 001 |
| P8 | PSD | Somatic Symptoms | 0.021 | -0.029 | 0.071 | 0.025 | 0.34 | 0.49 | 0.7 | .405 | .868 | .005 |

| | | | | | | | | | | | |
|---|---|---|---|---|---|---|---|---|---|---|---|
| Pz | PSD | Intercept | 0.156 | 0.053 | 0.260 | 0.052 | 3.60 | 0.40 | 9.0 | .003 | .021 | .059 |
| Pz | PSD | Bladder Pain | 0.002 | -0.005 | 0.009 | 0.004 | 0.17 | 0.40 | 0.4 | .518 | .974 | .003 |
| Pz | PSD | Menstrual Pain | 0.003 | -0.001 | 0.007 | 0.002 | 0.69 | 0.40 | 1.7 | .192 | .968 | .012 |
| Pz | PSD | Somatic Symptoms | -0.015 | -0.060 | 0.030 | 0.023 | 0.16 | 0.40 | 0.4 | .523 | .93 | .003 |
| T7 | PSD | Intercept | 0.258 | -0.018 | 0.534 | 0.140 | 9.79 | 2.86 | 3.4 | .066 | .236 | .023 |
| T7 | PSD | Bladder Pain | -0.003 | -0.022 | 0.015 | 0.009 | 0.38 | 2.86 | 0.1 | .718 | .974 | .001 |
| T7 | PSD | Menstrual Pain | 0.005 | -0.006 | 0.016 | 0.006 | 2.08 | 2.86 | 0.7 | .395 | .968 | .005 |
| T7 | PSD | Somatic Symptoms | -0.024 | -0.145 | 0.096 | 0.061 | 0.45 | 2.86 | 0.2 | .693 | .931 | .001 |
| T8 | PSD | Intercept | 0.050 | -0.120 | 0.220 | 0.086 | 0.37 | 1.08 | 0.3 | .562 | .701 | .002 |
| T8 | PSD | Bladder Pain | -0.004 | -0.016 | 0.007 | 0.006 | 0.61 | 1.08 | 0.6 | .455 | .97 | .004 |
| T8 | PSD | Menstrual Pain | 0.000 | -0.007 | 0.007 | 0.003 | 0.00 | 1.08 | 0.0 | .964 | .968 | < . 001 |
| T8 | PSD | Somatic Symptoms | -0.052 | -0.126 | 0.022 | 0.038 | 2.08 | 1.08 | 1.9 | .168 | .794 | .013 |
| TP10 | PSD | Intercept | 0.098 | -0.026 | 0.222 | 0.063 | 1.41 | 0.58 | 2.4 | .122 | .283 | .017 |
| TP10 | PSD | Bladder Pain | 0.000 | -0.008 | 0.009 | 0.004 | 0.00 | 0.58 | 0.0 | .934 | .974 | < . 001 |
| TP10 | PSD | Menstrual Pain | 0.001 | -0.004 | 0.006 | 0.003 | 0.14 | 0.58 | 0.2 | .625 | .968 | .002 |
| TP10 | PSD | Somatic Symptoms | -0.004 | -0.058 | 0.050 | 0.027 | 0.01 | 0.58 | 0.0 | .881 | .931 | < . 001 |
| TP9 | PSD | Intercept | -0.072 | -0.257 | 0.113 | 0.094 | 0.76 | 1.29 | 0.6 | .444 | .618 | .004 |
| TP9 | PSD | Bladder Pain | 0.000 | -0.012 | 0.013 | 0.006 | 0.00 | 1.29 | 0.0 | .956 | .974 | < . 001 |
| TP9 | PSD | Menstrual Pain | -0.003 | -0.010 | 0.005 | 0.004 | 0.76 | 1.29 | 0.6 | .443 | .968 | .004 |
| TP9 | PSD | Somatic Symptoms | 0.007 | -0.074 | 0.087 | 0.041 | 0.03 | 1.29 | 0.0 | .874 | .931 | < . 001 |

*Note.* Results for the Intercept model are equivalent across all electrodes and are therefore not presented for each individual electrode; *df* for the numerator and denominator were 1 and 143, respectively; LL = lower level; UL = upper level.
